# Modelling the impact of reducing control measures on the COVID-19 pandemic in a low transmission setting

**DOI:** 10.1101/2020.06.11.20127027

**Authors:** Nick Scott, Anna Palmer, Dominic Delport, Romesh Abeysuriya, Robyn Stuart, Cliff C. Kerr, Dina Mistry, Daniel J. Klein, Rachel Sacks-Davis, Katie Heath, Samuel Hainsworth, Alisa Pedrana, Mark Stoove, David P. Wilson, Margaret Hellard

## Abstract

**Aims:** We assessed COVID-19 epidemic risks associated with relaxing a set of physical distancing restrictions in the state of Victoria, Australia – a setting with low community transmission – in line with a national framework that aims to balance sequential policy relaxations with longer-term public health and economic need.

**Methods:** An agent-based model, *Covasim*, was calibrated to the local COVID-19 epidemiological and policy environment. Contact networks were modelled to capture transmission risks in households, schools and workplaces, and a variety of community spaces (e.g. public transport, parks, bars, cafes/restaurants) and activities (e.g. community or professional sports, large events). Policy changes that could prevent or reduce transmission in specific locations (e.g. opening/closing businesses) were modelled in the context of interventions that included testing, contact tracing (including via a smartphone app), and quarantine.

**Results:** Policy changes leading to the gathering of large, unstructured groups with unknown individuals (e.g. bars opening, increased public transport use) posed the greatest risk, while policy changes leading to smaller, structured gatherings with known individuals (e.g. small social gatherings) posed least risk. In the model, epidemic impact following some policy changes took more than two months to occur. Model outcomes support continuation of working from home policies to reduce public transport use, and risk mitigation strategies in the context of social venues opening, such as >30% population-uptake of a contact-tracing app, physical distancing policies within venues reducing transmissibility by >40%, or patron identification records being kept to enable >60% contact tracing.

**Conclusions:** In a low transmission setting, care should be taken to avoid lifting sequential COVID-19 policy restrictions within short time periods, as it could take more than two months to detect the consequences of any changes. These findings have implications for other settings with low community transmission where governments are beginning to lift restrictions.

## Introduction

Following a rise in COVID-19 cases, in March 2020 the Australian government introduced mandatory quarantine periods for people returning from overseas, as well as a variety of physical distancing policies, including closing pubs, bars, entertainment venues, churches/places of worship, restricting restaurants and cafes to take-away only, and limiting public gatherings to two people [1]. Two months after these policies were introduced, available epidemic data indicate that they were successful in disrupting the spread of COVID-19, with fewer than 55 cases per day diagnosed between 12 April and 8 May, down from a peak of 469 diagnosed cases on 28 March [2, 3]. Acknowledging that maintaining these restrictions for an extended period would be socially and economically unfeasible, the federal government released a framework for a *COVIDSAFE Australia* [4] on 8 May. This framework outlined a three-step sequence of policy options across the education, retail, hospitality, sport and health sectors that would enable their reopening under sustainable conditions, and allowed states and territories to adopt different policies at different times based on their specific COVID-19 conditions. The federal and jurisdictional governments also implemented public health measures to mitigate risks associated with relaxing policies, including a scale-up of testing capacity and the release of the *COVIDSafe* smartphone app, which records community contacts via Bluetooth connection to enable contact tracing in the event that someone with the app is diagnosed.

For countries entering COVID-19 response phases that involve relaxing restrictions, it is important to carefully consider the sequence and timing of relaxing policies so as not to compromise the overall effectiveness of the response. Each component that is relaxed will increase the risk of COVID-19 transmission, however it is currently unclear under which conditions the increased risk would be sufficient to allow an uncontrolled outbreak. Epidemic modelling is a crucial tool that can provide insight into the likely specific and combined impact of relaxing individual control measures, as well as the time required to monitor and observe the impact of relaxing these measures.

Epidemic models can be broadly classified as either population-level or individual-level. Population-level models are simpler and faster to produce, but they can only provide population-average estimates of the impact of policy changes. This is because they divide a population into a small number of discrete risk categories and assume homogeneous mixing and transmission risks within each category. In contrast, agent-based models use a set of autonomous ‘agents’ to represent a population and offer a more complex method for simulating individual-level characteristics and human behaviour [5]. In reality, the risk of COVID-19 transmission is highly heterogeneous and driven by the contact networks of individuals, which are dependent on age, household structure and participation in different social and community activities. Moreover, the impact of interventions to slow the spread ofbCOVID-19, such as contact tracing and quarantine measures, are highly contact network dependent. Interventions that are specific to contact networks can only be captured in individual-level models.

To our knowledge, no modelling is currently available for Australia that provides scenario analyses of the impact of “micro-policy” changes being proposed in the COVIDSAFE Australia framework. Population-level models [6–9] have been useful in informing a “flatten the curve” narrative and supporting the initial roll-out of physical distancing policies. Agent-based models are increasingly being used to simulate the impact of social distancing measures on COVID-19 transmission (either influenza models that have been modified [10–13], including for Australia [11, 14], or newly developed COVID-19 models [15–17]); however these models are currently only considering the implementation of contact tracing, quarantine or social distancing policies rather than their release.

In this study we used an agent-based model, *Covasim* [18], to assess the risks associated with relaxing various physical distancing and lockdown policies in Victoria, Australia. In this low transmission settings, we estimate how effective interventions need to be to mitigate these risks. Providing modelled estimates for these policy changes will be crucial, not just for Australia but globally in other settings with low transmission, and as countries begin to reopen.

## Methods

### Setting

We modelled the state of Victoria, Australia. Victoria is Australia’s second most populous state with an estimated population of 6.63 million (~26% of the nation’s total) [19], and age structure as shown in Appendix A, Figure S1. As of 17 May, Victoria had 1,561 confirmed COVID-19 cases, 835 (53%) of which were acquired through overseas travel, and 18 COVID-19-related deaths [20]. The Victorian epidemic has followed a similar trajectory to Australia as a whole; an increase in daily new diagnoses throughout March to a peak of 111 on 29 March, followed by a decline as various restrictions (outlined in Appendix B) were imposed. Throughout May there were two small “cluster” outbreaks detected at workplaces, where geographically isolated transmission occurred through known contacts leading to approximately 5-20 new diagnoses per day from 1 May to 17 May.

### Model overview

The *Covasim* model is described in detail on medrxiv [18] and has been applied to a number of high, middle and low-income settings, including a number of states in the USA and countries across Africa [21]. In brief, each person in the model is characterised by a set of demographic, disease and intervention status variables. Demographics variables include: age (one-year brackets); uniquely identified household contacts; uniquely identified school contacts (for people aged 5-18); uniquely identified work contacts (for people aged 18-65); and average number of daily community contacts in a collection of community networks and settings (described in the following sections). Disease variables include: infection status (susceptible, exposed, recovered or dead); viral load (time-varying); age-specific susceptibility (Table S2); and age-specific probabilities of being symptomatic, experiencing different disease severities (mild, severe, critical), and mortality (Table S2). Person-level intervention status variables include: diagnostic status (untested, tested and waiting for results, tested and received results) and quarantine status (yes/no).

Transmission is modelled to occur when a susceptible individual is in contact with an infectious individual through one of their contact networks. The per-day probability of transmission per contact with an infected person (“transmissibility”) is calibrated to match the epidemic dynamics observed, and is weighted according to whether the infectious individual has symptoms, and the type/setting of the contact (e.g. household contacts are more likely to result in transmission than community contacts).

### Contact networks

The model allows people to be a part of multiple independent contact networks. Within each network, a “contact” is a link between two people indicating that transmission would be possible if one of them were infected. The model is designed so that each individual can be a part of an arbitrary number of contact networks used to approximate transmission dynamics associated with different activities or specific public spaces. For this analysis, we considered networks and settings most likely to be subject to a policy change in Australia, with contact networks explicitly modelled for: households; schools; workplaces; social networks; cafés and restaurants; pubs and bars; public transport; places of worship; professional sport; community sport; beaches; entertainment (cinemas, performing arts venues etc); national parks; public parks; large events (concerts, festivals, sports games etc.); child care; and aged care.

Each contact network is defined by a set of properties: the percentage (and age range) of the population who are a part of it; the average number of contacts per day associated with these activities; whether the contacts are known or random; the type of network structure (random or cluster - for example public transport is random while schools/workplaces are clustered); the risk of transmission relative to a household contact (scaled to account for frequency of some activities); the effectiveness of contact tracing that might occur; and the effectiveness of quarantine at reducing transmission (e.g. quarantine may be effective for workplace transmission, not effective for household transmission, and partially effective for community transmission due to imperfect adherence).

Details of the contact networks are provided in Appendix D.

### Model initialization: household size and age structure

The model population was initialized through the generation of households. Individual households were explicitly modelled based on the households size distribution for Australia [22], with each person in the model assigned to a house. To assign people in the model an age, a single adult was selected for each household as an index, whose age was randomly sampled from a subset of the Victorian adult population (all adults 22 years and older and a percentage of 18-21 year olds - 20%, 40%, 60%, and 80% of people aged 18, 19, 20 and 21, respectively) to ensure that at least one adult was in each household. The age of additional household members was then assigned according to Australian age-specific household contact estimates (from Prem et al. [23], Figure S2), by drawing the age of the remaining members from a probability distribution based on the row corresponding to the age of the index member. The resulting age distribution of the model population, compared to the Victorian population, is provided in Figure S1.

### Other contact networks

School classrooms were explicitly modelled. Classroom sizes were drawn randomly from a Poisson distribution with mean 21, the Victorian average [24]. People in the model aged 5-18 years were assigned to classrooms with people of the same age. Each classroom had one randomly selected adult (>21 years) assigned to it as a teacher. The school contact network was then created as a collection of disjoint, completely connected clusters (i.e. classrooms).

Similarly, a work contact network was created as a collection of disjoint, completely connected clusters of people aged 18-65 years. The size of each cluster was drawn randomly from a Poisson distribution with mean equal to the estimated average number of daily work contacts (Table S4). Other clustered contact networks, such as places of worship, community sports, professional sports, child care and aged care were generated analogously (with transmissibility scaled to account for event frequency; Appendix D).

Random contact networks (e.g. public transport) were generated by allocating each person a number of contacts drawn from a Poisson distribution with mean as per Appendix D. Unlike the clustered contact networks, the contacts in random contact networks were resampled at each time step in the model (representing days).

### Modelling interventions and policy changes

Policy scenarios modelled were informed by the COVID-19 public health response in Victoria [1] and the *COVIDSAFE Australia* framework [4], and included scenarios related to: the effectiveness of contact tracing; compliance with physical distancing; restricting access to hospitality and entertainment venues and other public spaces; restricting access to places of worship; restricting the size of social gathering; restricting community and professional sport; closing schools and childcare settings; closing non-essential workplaces, retail outlets and health care; and restricted travel across jurisdictional borders and domestic travel.

Each policy change is linked to one or more networks, and can potentially influence the whole population. For example, if non-essential work begins, this would increase the size of the work network, as well as increasing transmissibility in public transport. See Appendix E for full list of modelled scenarios.

### Model parameters

Epidemiological data for the daily number of tests conducted, new diagnoses and new severe cases, critical cases and deaths was obtained from the Victorian Department of Health [20, 25]. Newly diagnosed cases were classified as “imported” to Victoria if their mode of acquisition was listed as travel overseas.

Disease specific parameters, including duration of incubation, infectious and symptomatic periods, and age-specific risks associated with disease severity and outcomes, were based on global published estimates (Table S1 and Table S2).

Parameters for contact networks and the effect of policy changes were obtained from a combination of the literature and a modified Delphi process (Appendix D). The modified Delphi process involved creation of a panel of 12 experts (a mixture of modellers, epidemiologists, qualitative researchers, social network researchers, infectious disease physicians and public health physicians), who participated in a video conference where they were introduced to the model and the interpretation of parameters. Panel members were then asked to make independent estimates of unknown parameters, which were collated and de-identified by the study team, and the median and range of each parameter was extracted. A follow-up video conference was held where the panel discussed the results and uncertainties and were provided an opportunity to revise any estimates. The distribution of responses for each parameter, as well as the final parameters used, are provided in Appendix D.

### Baseline scenario and calibration

A baseline scenario was run between 1 March and 30 April, including with the policy changes that had occurred over that period, and the parameter for the overall probability of transmission per contact was calibrated such that the model projections fit the data on number of diagnoses and deaths.

### Scenario set 1: Policy relaxations

Multiple scenarios were run, in which different policy restrictions were lifted in isolation starting from 15 May (the date of analysis): opening pubs/bars; allowing large events; opening cafes and restaurants; allowing community sports; allowing small social gatherings; opening entertainment venues (e.g. cinemas, performing arts); removing work from home directives (resulting in greater public transport use as well as more work interactions); and opening schools. The parameter and network configuration changes associated with relaxing each restriction are described in Appendix D. For each scenario, a number of new infections were introduced for modelling purposes (a theoretical five infections on 15 May) to restart the epidemic and test the robustness of the new policy configuration to outbreaks.

### Scenario set 2: Contact tracing smartphone app

We estimated the threshold population-level coverage that a contact tracing smartphone app (i.e. *COVIDSafe*) would need to mitigate the risks of relaxing different policies. The threshold target was calculated to mitigate the risks associated with the policies of opening of pubs/bars and removing work from home directions, as these were the policies found to have the greatest risk (see results). Multiple scenarios were run where these policies were changed but with population-level coverage of the contact tracing app ranging from 0-50%.

### Scenario set 3: Physical distancing policies within venues

Policy options are available and being utilized by governments mitigate the risks associated with opening of cafés, restaurants, pubs and bars; for example, transmissibility in these settings could be reduced by implementing the “4 square metre rule” and similar limits on customer numbers, or restricting venues to outside service only. We estimate how effective these additional interventions would need to be to mitigate the risks associated with opening these venues. Opening pubs/bars was used as an example as it was found to pose the greatest risk, and multiple scenarios were run where transmissibility within pubs and bars was reduced by 0-50%.

### Scenario set 4: Patron records at venues

An additional policy option being used is for venues (pubs/bars/cafes/restaurants) to keep mandatory identification records of patrons, which would enable contact tracing following a diagnosed case (similar to a contact tracing smartphone app, but with much higher coverage over a smaller time and place). We estimate the threshold compliance with this policy required to mitigate the risks associated with opening these venues. Multiple scenarios were run where pubs/bars were opened but with the capacity to contact trace 40-80% of contacts following a within-venue transmission event.

## Results

### Model calibration

A reasonable model fit was obtained (Figure 1) that included the initial increase in cases observed followed by the subsequent decline in cases following the introduction of specific policy changes. We estimate that by April 30, approximately 2000 people had been infected with COVID-19, of which approximately 1600 (80%) had been diagnosed. The undiagnosed proportion primarily includes asymptomatic cases.

**Figure 1:**
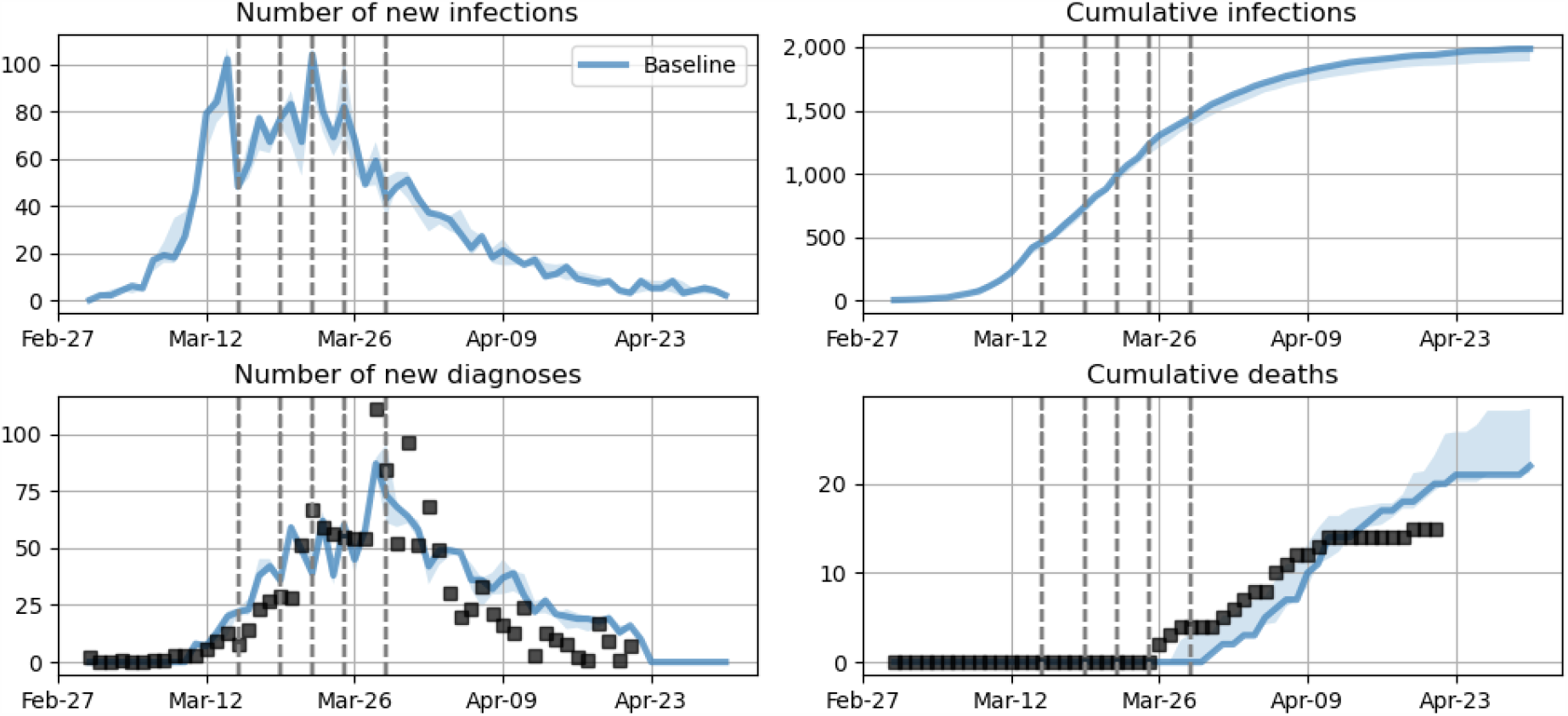
Model calibration and baseline projection for the initial epidemic wave in Victoria. The probability of transmission per contact was varied such that the model fit the observed number of diagnoses and deaths over time. Baseline projections (blue) include policy changes that occurred on March 19, 21, 22 and 29 (dashed vertical lines, described in detail in Appendix D).

### Scenario set 1: Policy relaxations

To estimate which policy change is associated with the greatest or quickest increase in infections we assessed the impact of lifting individual policies separately, on May 15^th^. The greatest risk of a rebound in cases comes from policy changes that facilitate random, once-off mixing in the community, or situations where individuals have a large number of contacts, particularly those that are unknown. This includes opening pubs and bars (without additional restrictions), removing work from home directives (which increases public transport and work interactions) or allowing large events (concerts, sporting crowds, protest marches). The least risk comes from policy changes that facilitate smaller numbers of contacts, or repeated contacts with the same people (e.g. small social gatherings) (Figure 2).

**Figure 2:**
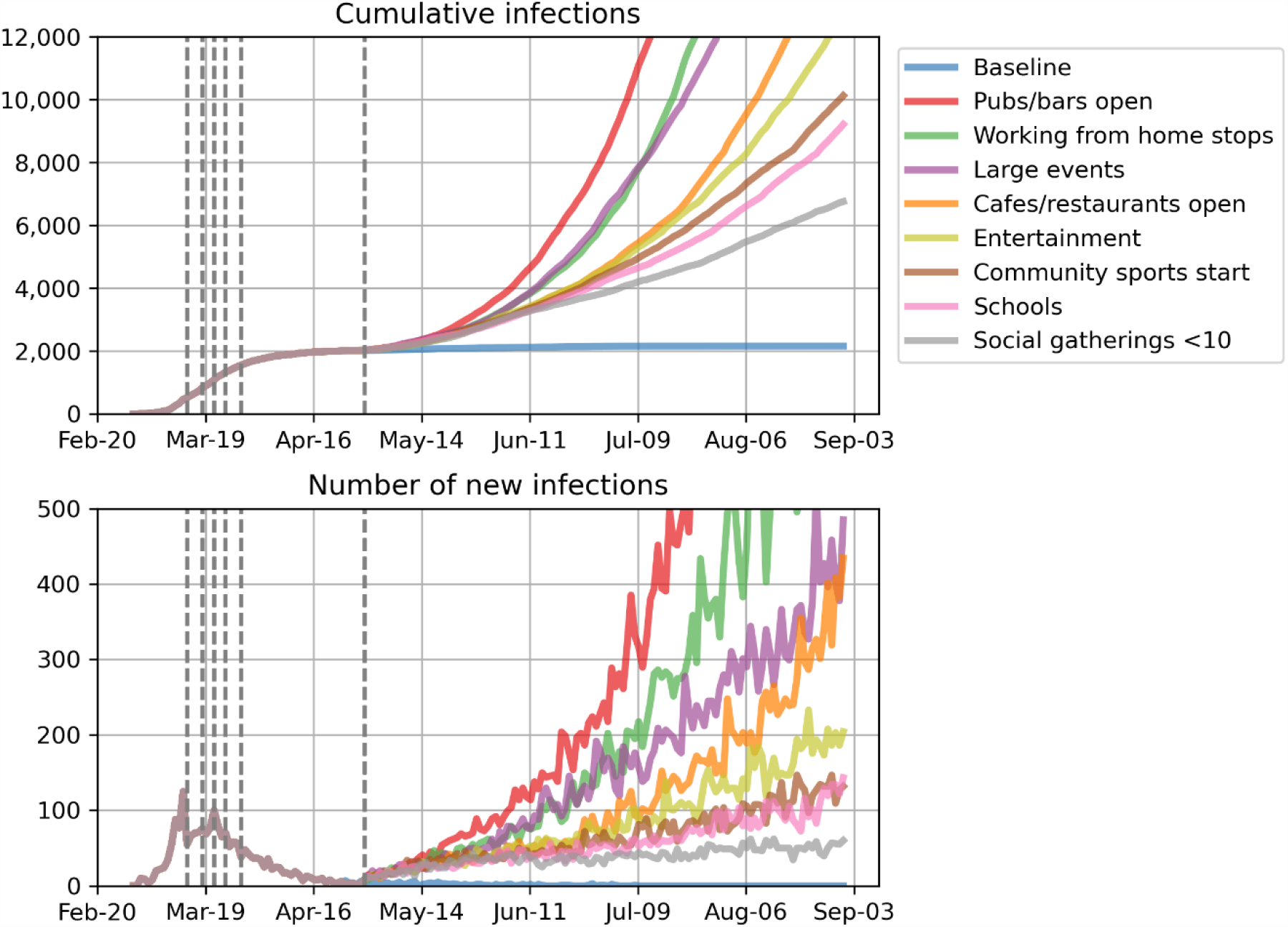
Impact of policy changes. Projected cumulative population-level infections when different policy restrictions are lifted. Dashed vertical lines show the dates of policy changes. In these projections, venues are modelled as being opened without additional physical distancing restrictions, and population-level coverage of the contact tracing smartphone app was set to 5% (estimated coverage at 15 May).

Importantly, for some policy changes the time before new infections begin to rapidly increase could be greater than two months (Figure 2, for example cafes/restaurants or entertainment venues opening).

### Scenario set 2: Contact tracing smartphone app

To assess the potential impact of the contact tracing smartphone app on mitigating an increase in new infections, we modelled two highest risk policy changes – the re-opening of pubs and bars and removing working from home directives – with varying degrees of app coverage. Greater than 30% coverage was required before the app showed significant impact on mitigating population-level transmission risks (Figure 3 for pubs and bars being opened, and Figure S5 for working from home directives being removed).

**Figure 3:**
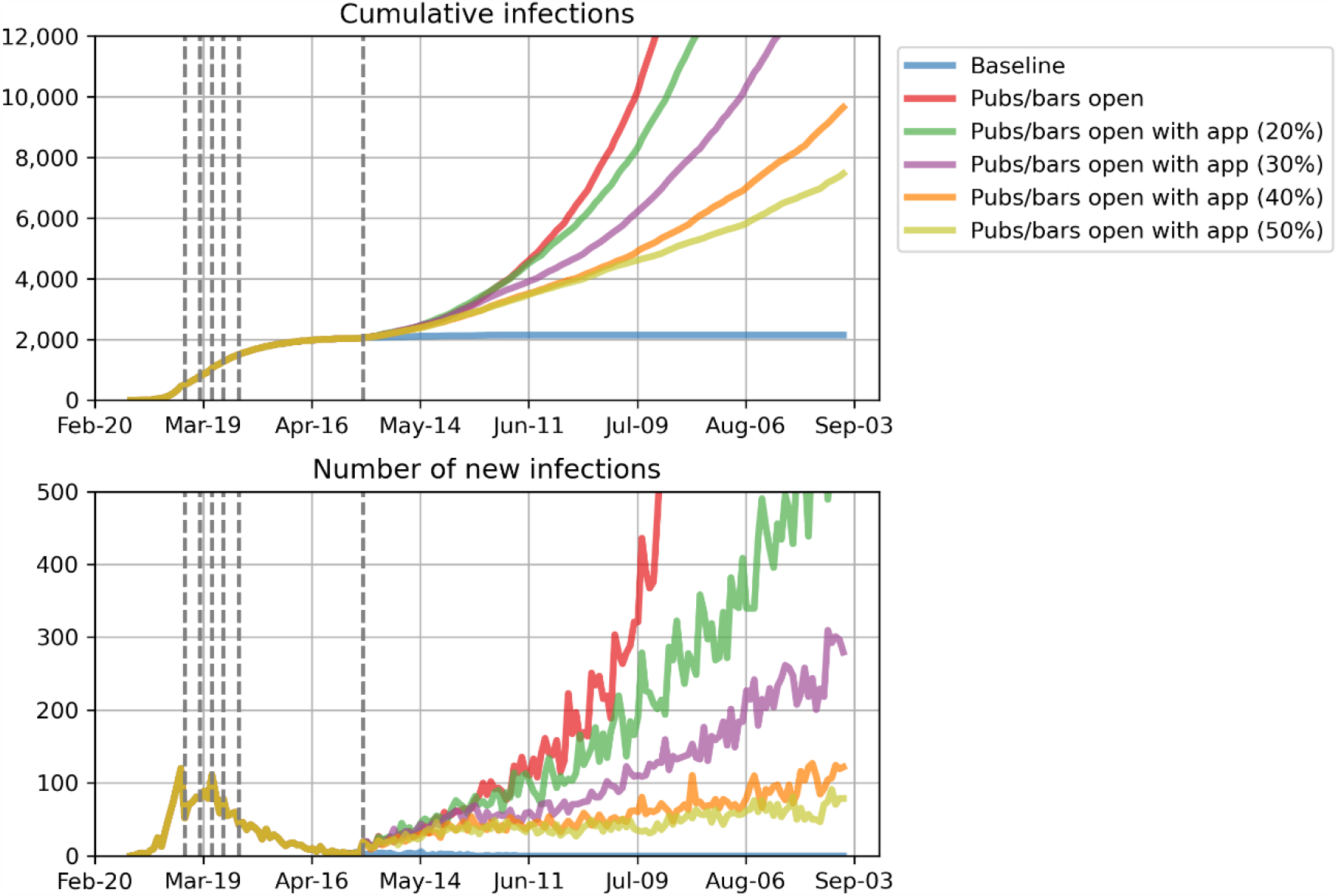
Impact of contact tracing smartphone app. Projected cumulative population-level infections when pubs and bars are opened, with different uptake of the smartphone app. Dashed vertical lines show the dates of policy changes.

### Scenario sets 3-4: Mitigation strategies in venues

Opening pubs and bars (without additional restrictions) was found to be the policy that led to the greatest increase in new infections. However, the model suggests that if physical distancing policies within these settings could reduce transmissibility by more than 40% they could considerably mitigate the risks of them opening (Figure 4). Alternatively, recording the identification of patrons attending pubs and bars to enable effective contact tracing would be an effective policy at a population-level if compliance was greater than 60% (Figure S6).

**Figure 4:**
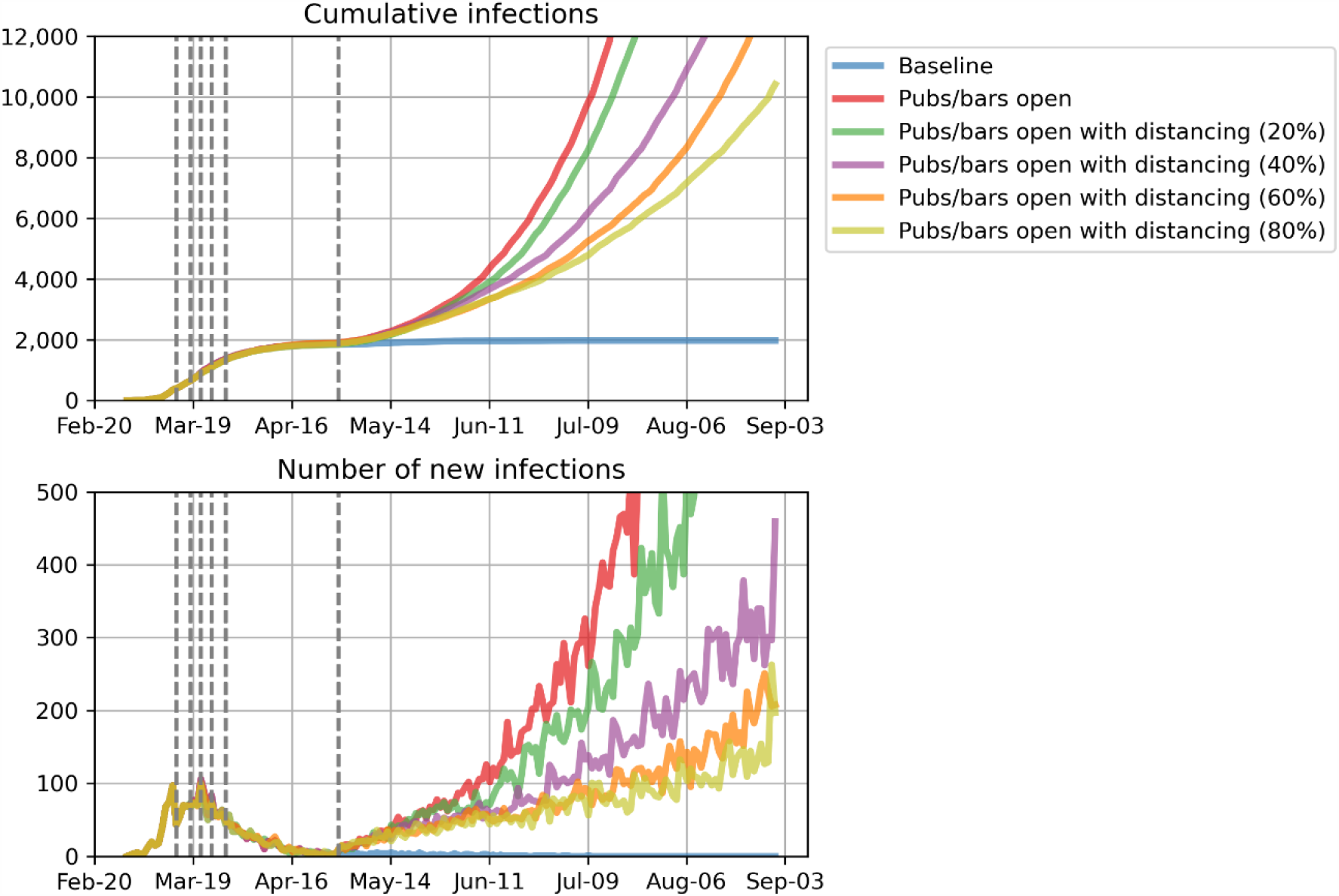
Impact of physical distancing policies combined with opening of pubs and bars. Projected cumulative population-level infections when pubs and bars are opened, with physical distancing policies (e.g. the “4 square metre rule”) that reduce transmissibility by 20-80%. Dashed vertical lines show the dates of policy changes. Population-level coverage of the contact tracing smartphone app was set to 5% (estimated coverage at 15 May).

## Discussion

Using an agent-based model we have simulated the relaxation of a variety of policy restrictions in a low transmission setting in Australia. We found that policy changes leading to large, unstructured contact networks (e.g. pubs and bars opening, increased public transport use through removal of work from home directives, or large events) posed the greatest risk, while policy changes leading to smaller, structured contact networks with known individuals (e.g. small social gatherings) posed the least risk. Importantly, the model suggests that it could take more than two months to detect increases in new infections from a change in policy, and therefore care should be taken in introducing multiple policy changes within short time periods. These outcomes, and this modelling tool, have implications for other settings with low community transmission where governments are lifting restrictions following relatively successful early responses.

Despite social and economic pressures to fast-track a return to normal conditions, our results suggest that restraint is needed, even in low transmission settings, because a resurgence in the epidemic following some policy changes could take more than two months to establish and be detected. In the model, contact tracing is effective for known contacts (Table S7); however, transmission to unknown community contacts can still occur, with frequency dependent on the mixing allowed by the policy environment. Transmission to an unknown contact would only be detected through their eventual symptomatic testing, allowing opportunity for them to further transmit during their infectious/pre-symptomatic stage. As with the index case, some of these additional infections will be of known contacts and effectively traced and isolated, however it is the chains of transmission through unknown contacts that may represent a minority of new cases initially, but if allowed to continue provide an increasing cumulative risk for epidemic expansion. It is therefore essential that accessible and efficient testing sites are available to complement contract tracing programs and detect community transmission from unknown sources. Once community transmission is detected, interventions to suppress a second epidemic wave would be required, for example geographically targeted testing or re-introduction of restrictions, to reverse the increasing trends projected in our scenarios. These interventions have not been modelled in this study and further work is required to assess how outbreaks can be managed without needing a complete policy reversal.

The greatest risks of a resurgence in cases were associated with policy changes that allowed individuals to have large contact networks (e.g. crowded public transport, crowded pubs/bars, sports events) that introduce once-off mixing between unknown individuals in the community. In particular, these findings support the Victorian government’s decision to extend work from home directions for people who are able until at least July 2020, to minimise use of public transport [1]. Further modelling work could assess whether staggered work starting times (to limit the number of riders), increased ventilation and cleaning, or face mask recommendations could mitigate the risks associated with increased public transport use. Policies creating large networks with random mixing were also found to be high risk even when they only affected a small subset of the population. For example, even though a minority of the population attend sporting events regularly, the random mixing of large numbers of contacts at a sporting event creates connectivity between smaller, clustered networks of known contacts, such as workplaces and households.

The lowest risks were associated with policy changes that led to smaller numbers of contacts for individuals, introduced organized contact network structure (e.g. known contacts), or introduced easily traceable contacts (e.g. family or small social gatherings). Under these network configurations, population-wide connectivity remains restricted, limiting the potential for wide-scale population spread. In addition, known contacts have a greater probability of being traced in a timely way when transmission does occur. These findings suggest that a smaller number of high intensity contacts is favourable compared to a large number of low intensity random contacts.

We found that a contact tracing smartphone app (i.e. *COVIDSafe*) would need greater than 30% effective population coverage to mitigate the risks associated with most policy relaxations. The effectiveness of the app relies on both the infected and susceptible person having the app and using it correctly (i.e. having their Bluetooth switched on and the app enabled), which means that if 30% of the population in a given time and place have downloaded the app, this would produce at most an additional 9% (30%*30%) of contacts able to be reliably traced. Importantly, the app also relies on a well-resourced, timely contact tracing and testing system to be in place to follow-up people identified as potentially exposed. As of end May, the COVIDSafe app had been downloaded by approximately 6 million Australians. This represents approximately 24% of the population, meaning that the app could trace at most an additional ~6% (24%*24%) of contacts. Therefore, while the app could be effective at high coverage, it is likely to have minimal impact for low-moderate coverage.

Based on the current epidemiological situation, we estimated that to mitigate the risks of opening pubs and bars (the policy change found to pose the greatest risk), physical distancing strategies that can reduce COVID-19 transmissibility by at least 40% in these settings are required. The model cannot identify what interventions may be able to achieve this, but this provides a useful target for designing interventions that consist of a mix of hygiene measures, physical distancing and limits to patron numbers. An additional policy option being implemented is for pubs and bars to keep mandatory identification records of patrons, which would enable rapid contact tracing following a diagnosed case. The model identified that this could be an effective policy if it enabled greater that 60% of contacts to be traced if an infected person was found to have attended a venue (Figure S5). Note that for mandatory identification to be as effective as the smartphone app, it needs to be more stringent, since the app has additional benefits by tracing multiple generations of transmissions rather than only those in the source setting.

Schools have been a topic of considerable policy discussion in Australia and internationally. Data are extremely limited on the susceptibility and transmissibility of COVID-19 among children, as well as health outcomes associated with infection. In our projections, opening schools was not predicted to lead to major population-level epidemic rebound, however it is critical to understand that this conclusion is based on some input parameters for which there is limited evidence. First, people aged under 20 years were assumed to be less susceptible to infection than people over 20 years (people aged 0-9 or 10-19 have relative susceptibility of 0.34 or 0.67 respectively, Table S2). Second, the probability of people under 20 years being symptomatic was lower than for people over 20 years, with asymptomatic cases having reduced transmissibility in the model. Third, the network structure for schools was clustered into classrooms with no links connecting different classrooms in a school, allowing more effective contact tracing following any outbreak. While this is the best available evidence at the time of writing, ongoing work is required to verify this preliminary data, and alternate inputs associated with COVID-19 dynamics and children would lead to different outcomes.

### Limitations and further work

The main limitations to this work are around model features, disease epidemiology parameters and contact network parameters.

This model currently only attributes basic properties to individuals, specifically age, household structure and participation in different contact networks. Therefore, the model does not account for any other demographic and health characteristics such as socioeconomic status, comorbidities (e.g. non-communicable diseases) and risk factors (e.g. smoking) and so cannot account for differences in transmission risks, testing, quarantine adherence or disease outcomes for different population subgroups. Further work is required with the specific aims of assessing the impact of policy changes on different subsets of the community. The model also does not include a geospatial component and so cannot capture geographic clustering of infections or concentration of interventions, including differential tracing app uptake in urban versus rural settings or among people attending particular events or settings, or concentrated testing in response to a localised outbreak. This means that our projections may be overestimating outbreak sizes as geographic clustering may slow epidemic spread.

Data reported on disease parameters such as duration of asymptomatic and infectious periods, as well as age-specific estimates of susceptibility, transmissibility and disease severity and are likely to be influenced by differences in surveillance systems in the countries they are being reported from. We have taken the best available data at the time, but this is likely to change as new information becomes available, and the model should be updated accordingly.

Contact networks are the most important factor driving COVID-19 transmission yet limited studies are available that provide the parameters needed to model them. The modified Delphi process used has potential biases in the non-randomly selected panel, and the large variation in parameter estimates suggests a high degree of uncertainty in contact network parameters. Despite this uncertainty, we argue that it is still important to consider these contact networks and the impact of policy changes on them. For example, studies are not available to quantify the relative transmissibility among public transport contacts compared to household contacts. However, omitting this parametrisation would implicitly either ignore public transport contacts, or assume that they are equal to household contacts. In this study we have instead assumed that they fall somewhere in between, but we do not know where and hence have used a panel to estimate. Similarly, if people are instructed to work from home, then the transmission risk on public transport would be expected to decrease. While the actual reduction is unclear, if this feature were not included then this would implicitly assume that there was no change. It is critical that these parameters are continually updated as new evidence becomes available, and that the model is used to compare multiple policy options rather than to directly estimate the effects of individual policies. In particular, conclusions should be drawn that relate to the network properties, rather than the policies; for example, statements about random contacts being worse than clustered contacts [26], rather than statements about specific policies.

## Conclusions

In settings with low community transmission, care should be taken to avoid introducing multiple policy changes within short time periods, as it could take greater that two months to detect the consequences of any changes. When selecting which policies to relax, the greatest risks are associated with large gatherings of people who do not know each other. In particular, working from home directions should be maintained to minimise use of public transport. Opening pubs/bars was identified as the greatest risk in Victoria; however these risks could be mitigated if either greater than 30% population-level coverage of a contact tracing smartphone app was achieved, transmissibility within venues was reduced by more than 40% through physical distancing policies, or patron identification records were kept that enabled greater that 60% of contacts to be traced if an infected person was found to have attended a venue.

## Data Availability

All parameter inputs used for this modelling study are available in the supplementary material.

## Acknowledgements

The authors would like to thank Allan J. Saul, Angela Davis, Joseph Doyle, Sherrie Kelly, Suman Majumdar for contributions to parameter estimates, and additional members of the Institute for Disease Modelling team who contributed to the base Covasim model. The authors gratefully acknowledge the support provided to the Burnet Institute by the Victorian Government Operational Infrastructure Support Program. RSD, MS and MH are the recipient of an Australian National Health and Medical Research Council fellowships.

## SUPPLEMENTARY MATERIAL

### APPENDIX A: Additional figures

**Figure S1:**
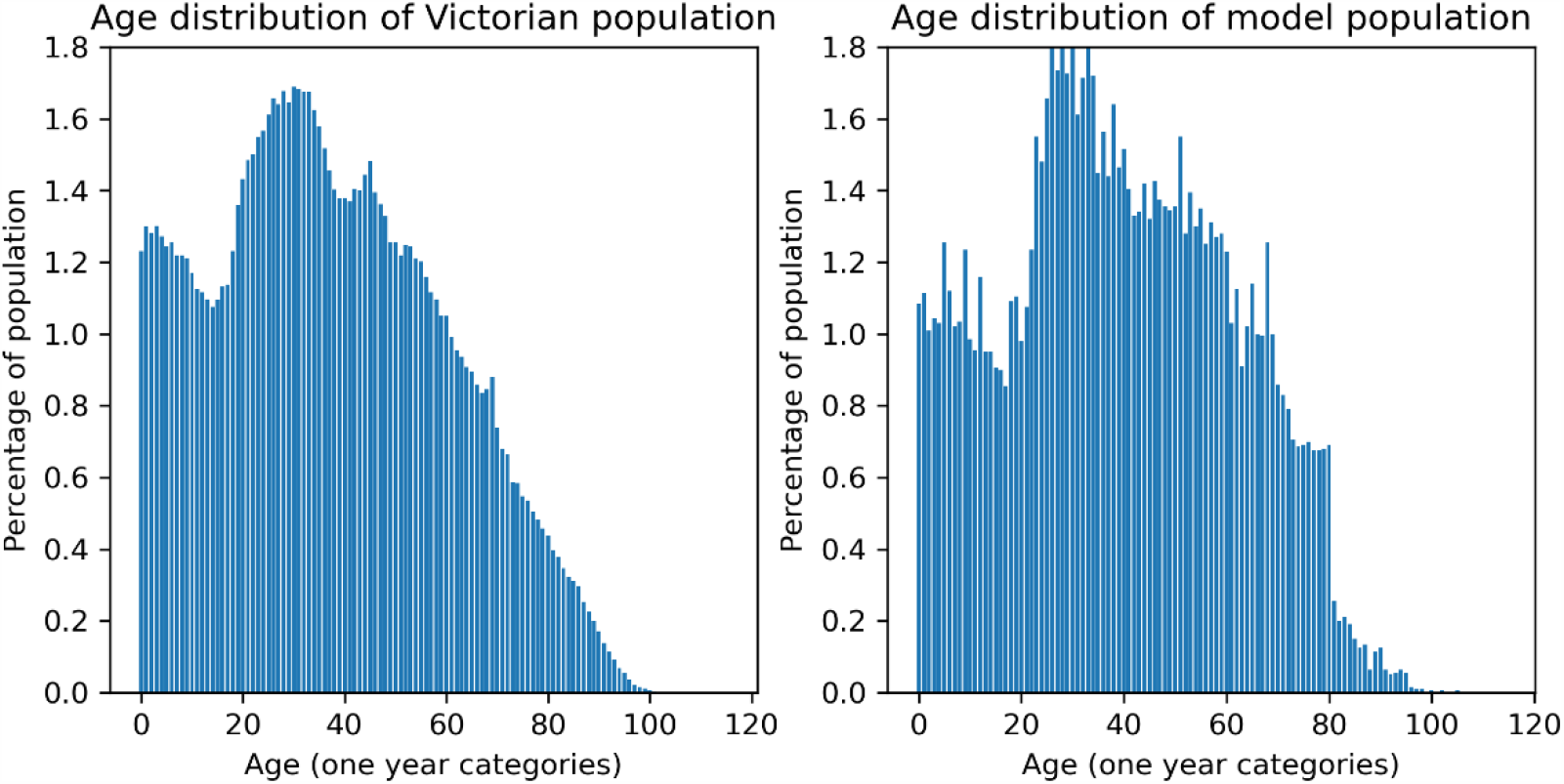
Age distribution (input vs modelled).

**Figure S2:**
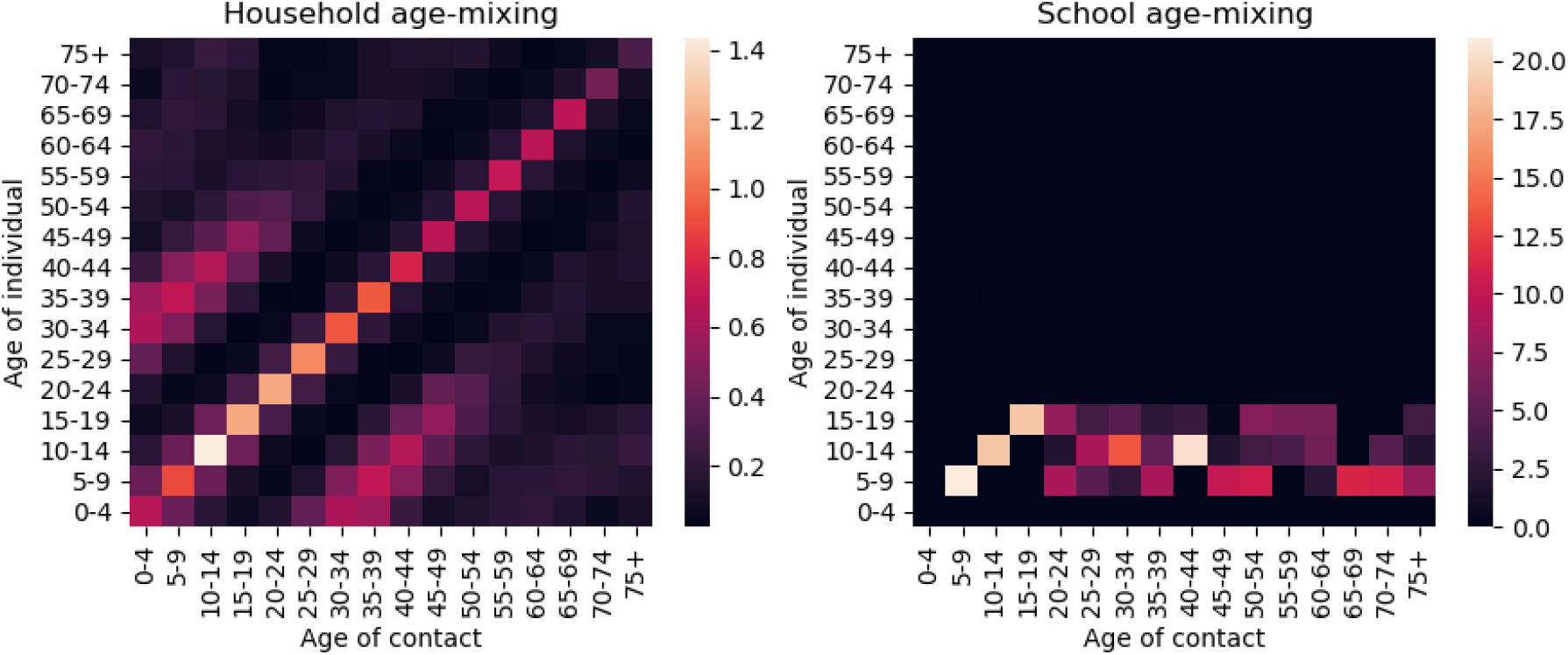
Age mixing within households and schools. Right: household mixing reproduced from Prem et al. [1] estimates). Left: within schools, students aged 5-18 were in classrooms with others of the same age, and one teacher per classroom (the asymmetry is due to all students having an adult teacher contact, but not all adults being teachers and having school contacts). The y-axis represents the age of the individual and the x-axis represents the age of their contacts. The colour represents average number of daily contacts.

**Figure S3:**
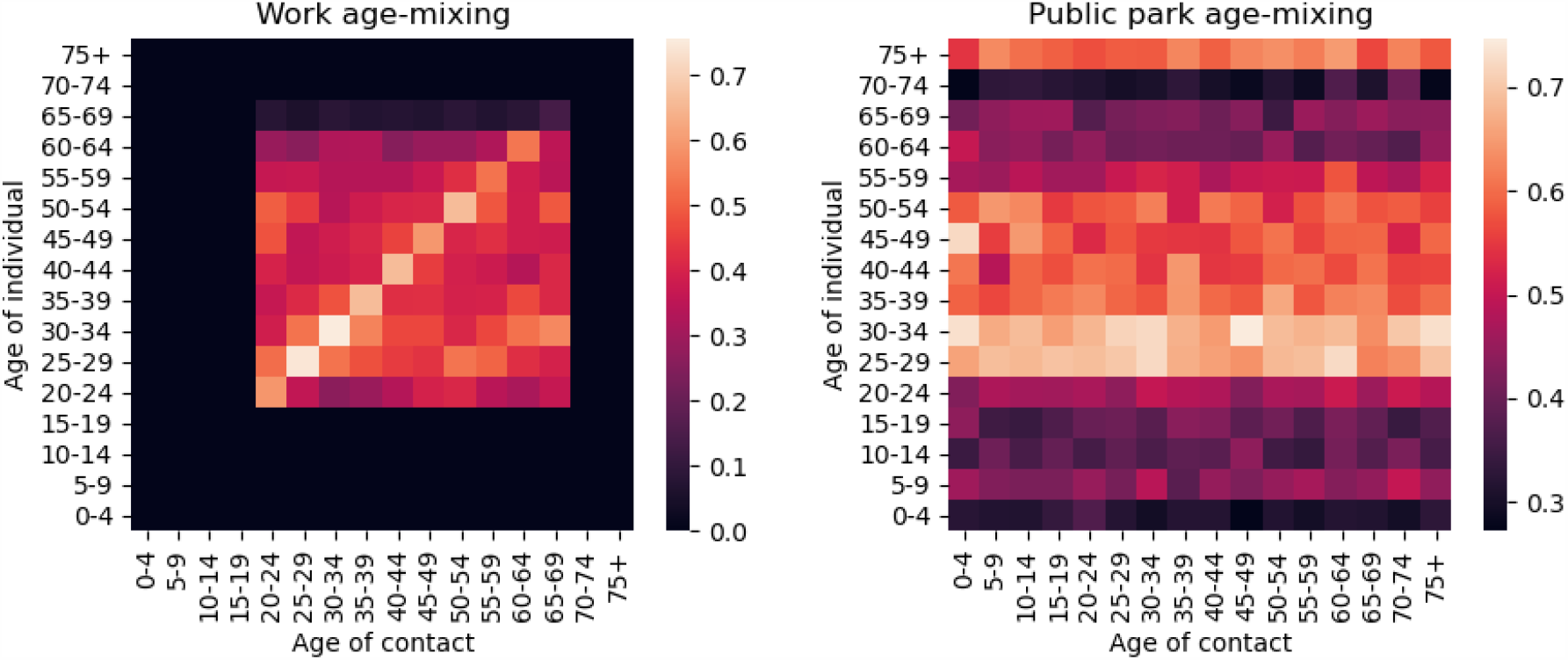
Examples of age-mixing within workplaces and public spaces. Left: at workplaces, adults aged 18-65 could mix with adults of any other age. The extra intensity on the 25-35-year-old diagonal is due to the disproportionate population age distribution in Victoria. Right: in public spaces, all ages could mix together. The y-axis represents the age of the individual and the x-axis represents the age of their contacts. The colour represents average number of daily contacts.

**Figure S4:**
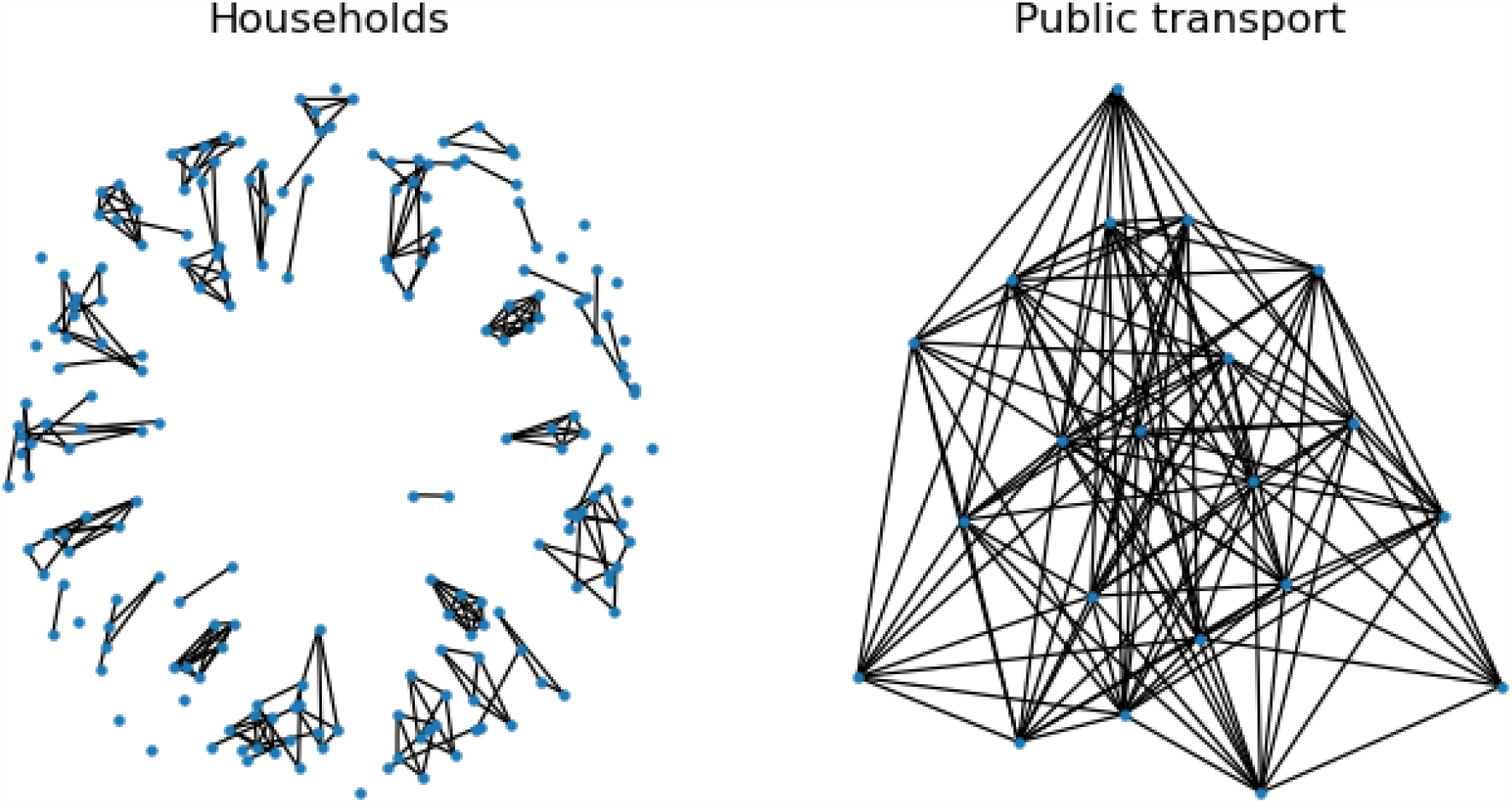
Example contact network structures between in the model. Left: the workplace network was modelled as clusters with size drawn from a Poisson distribution, and was fixed throughout a simulation. Right: some the community transmission networks, such as public transport, were modelled such that each individual had a number of contacts that were randomly assigned, and were re-assigned each day.

**Figure S5:**
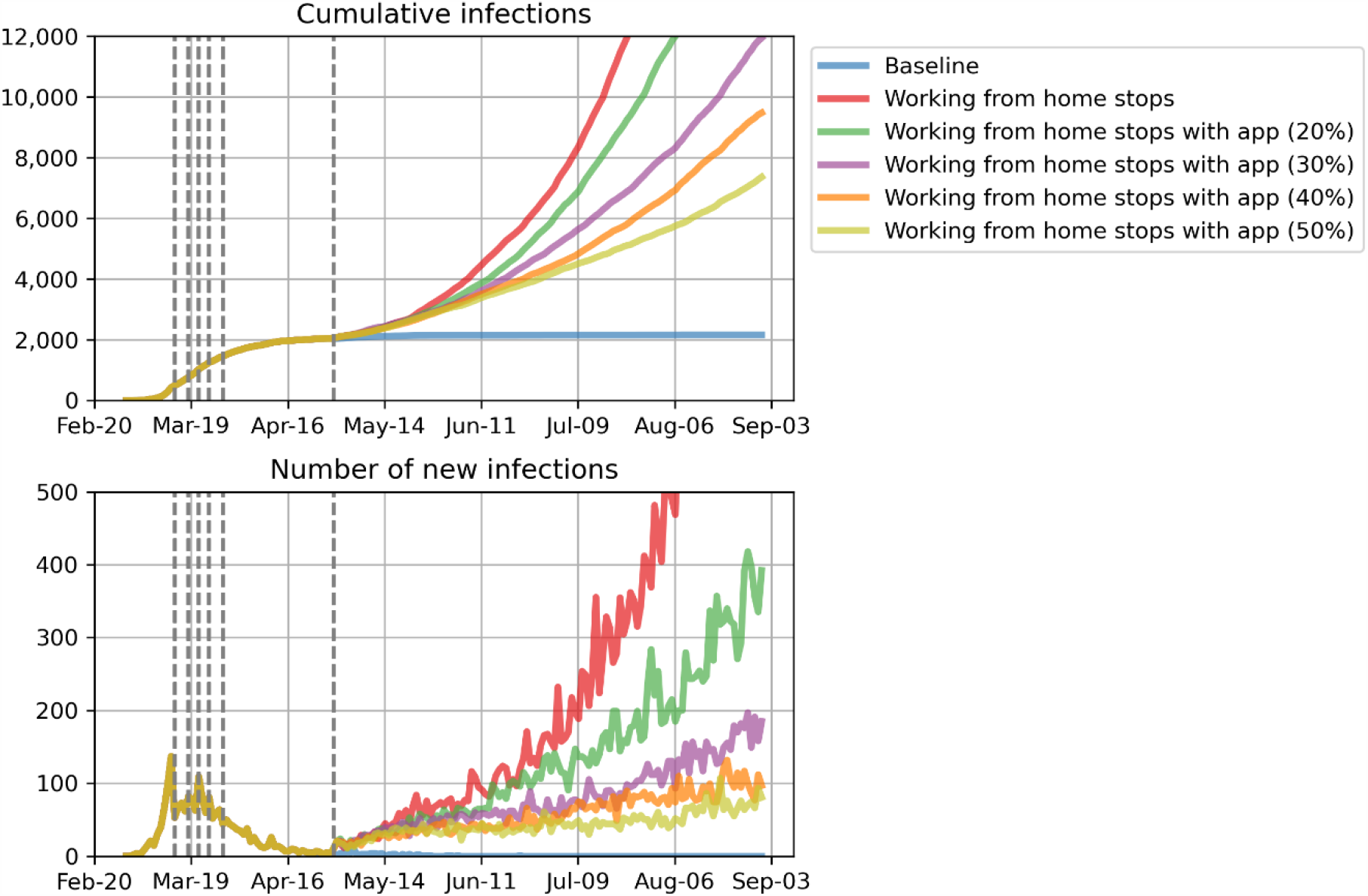
Impact of contact tracing smartphone app. Projected cumulative population-level infections when work from home directives are removed, with different uptake of the smartphone app. Dashed lines show the dates of policy changes.

**Figure S6:**
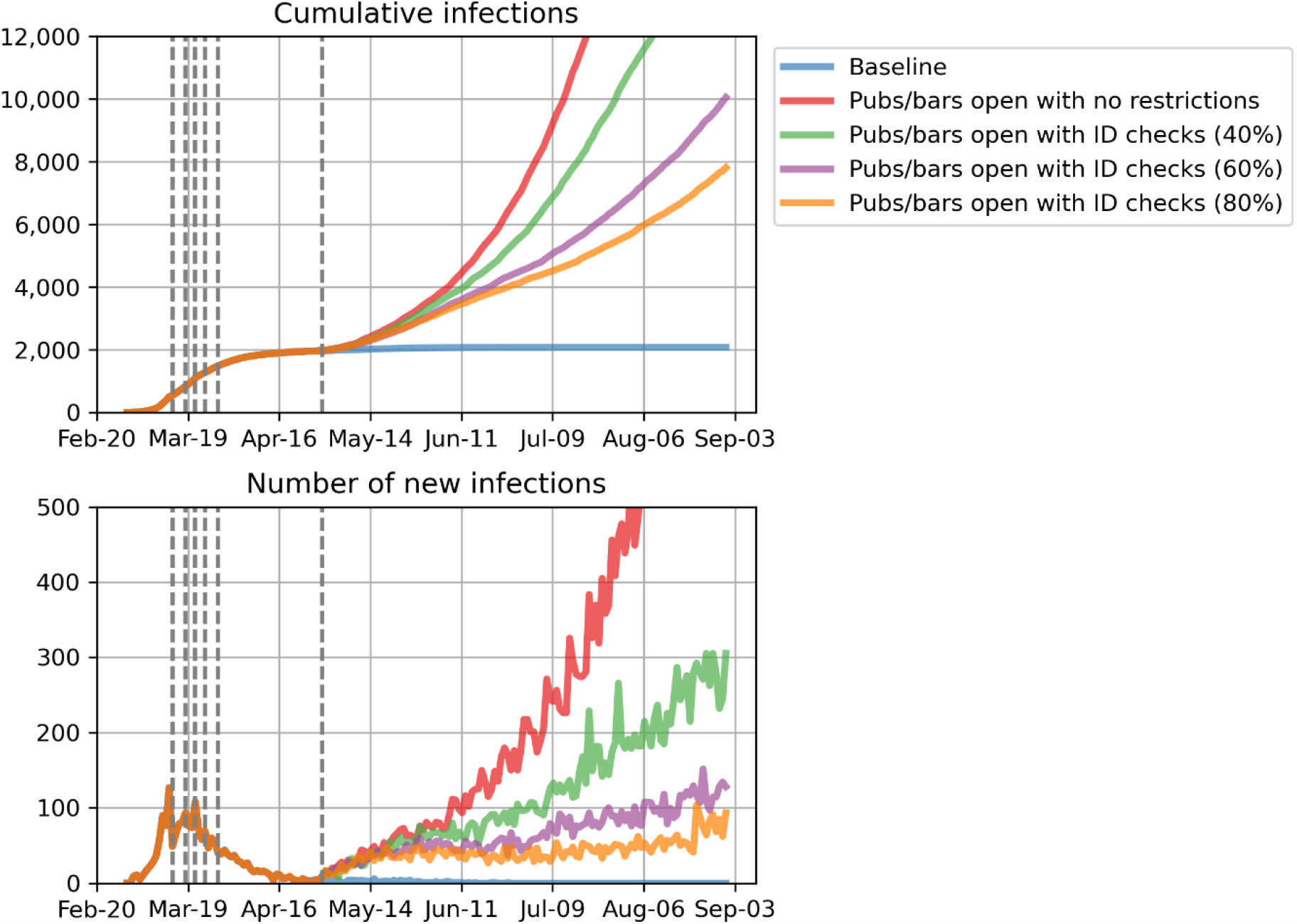
Impact of identification collection alongside the opening of pubs and bars. Projected cumulative population-level infections when pubs and bars are opened, with compulsory identification recording enabling 40-80% of contacts from those venues to be traced within one day of a diagnosed case. Dashed lines show the dates of policy changes. Population-level coverage of the contact tracing smartphone app was set to 5% (estimated coverage at 15 May).

**Figure S7:**
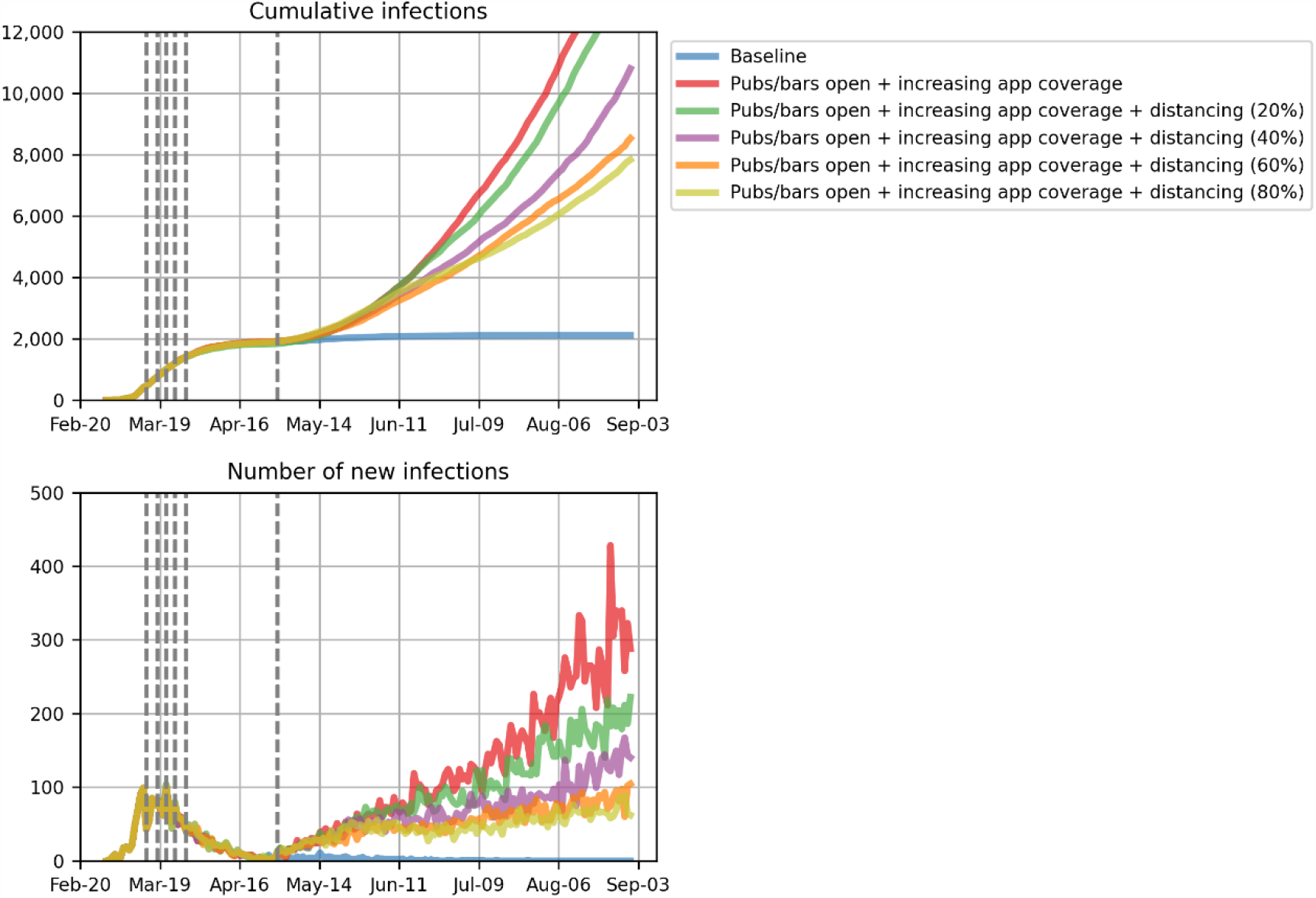
Impact of physical distancing policies in pubs and bars <u>combined with smartphone app coverage scale-up to 25% by 15 June</u>. Projected cumulative population-level infections when pubs and bars are opened, with compulsory identification recording enabling 40-80% of contacts from those venues to be traced within one day of a diagnosed case. Dashed lines show the dates of policy changes.

### APPENDIX B: Policy changes occurring in Victoria, Australia

Summarized from Wikipedia [2].

- 1 Feb: Travel restrictions from China
- 1 Mar: Travel restrictions from Iran
- 5 Mar: travel restrictions from South Korea
- 11 Mar: travel restrictions from Italy
- 15 Mar: gatherings of more than 500 people cancelled
- 15 Mar: all international travellers must self-isolate for 14 days
- 19 Mar: indoor gatherings limited to 100 people
- 20 Mar: Australia closes borders to all non-residents and non-Australian citizens
- 21 Mar: 4 square metre social distancing rule for people in any enclosed spaces
- 22 Mar: pubs, bars, entertainment venues, cafes, cinemas, restaurants, places of worship closed (or take-away only)
- 29 Mar: public gatherings limited to two people.
- 29 Mar: People over 70 years, people with chronic illness over 60 years, or Indigenous Australians over 50 urged to self-isolate
- 29 Mar: only four reasons to leave home: shopping for essentials; for medical or compassionate needs; exercise in compliance with the public gathering restriction of two people; and for work or education purposes

**Figure S8:**
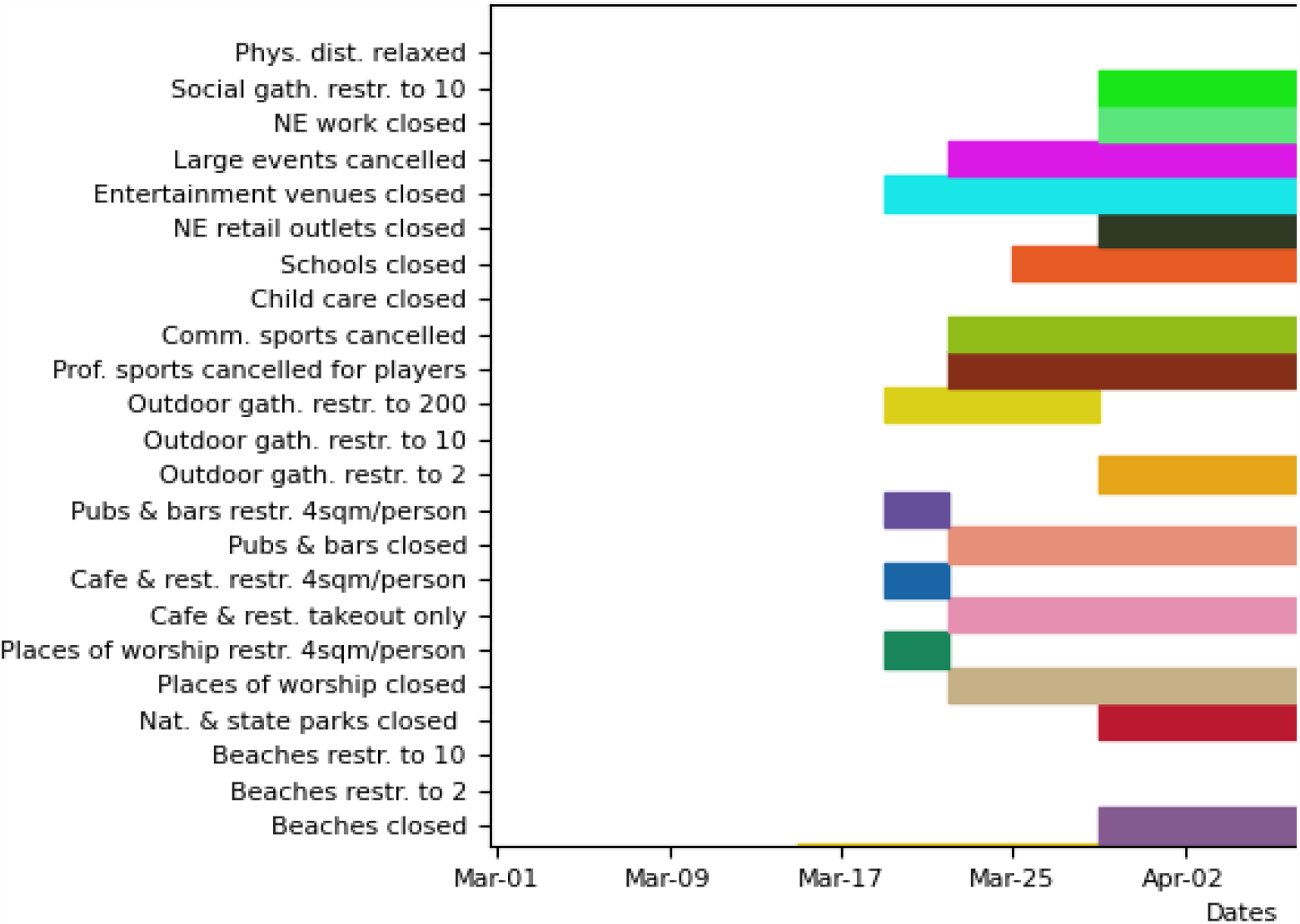
Policy changes and restrictions that were implemented in the model.

### APPENDIX C: Model parameters

**Table S1:** model parameters.

| Description | Value | Source |
| --- | --- | --- |
| Disease-related parameters | Distribution (mean, std) |  |
| Period from exposure to infectiousness | Lognormal(4.6,4.8) | From Lauer et al., 2020 [3]; additional sources Du et al., 2020 [4]; Nishiura et al., 2020 [5]; Pung et al., 2020 [6] |
| Period from infectious to symptomatic | Lognormal(1,1) | He et al., 2020 [7] report that infectiousness started from 2.3 days (95% CI, 0.8–3.0 days) before symptom onset and peaked at 0.7 days (95% CI, –0.2–2.0 days) before symptom onset. Gatto et al., 2020 [8] estimate a pre-symptomatic period of 1.3 days. |
| Duration for asymptomatics to recover | Lognormal(8,2) | Wolfel et al., 2020 [9] |
| Duration for mild symptoms to recover | Lognormal(8,2) | Wolfel et al. [9] |
| Duration for severe symptoms to recover | Lognormal(14,2.4) | Verity et al. [10] |
| Duration for critical symptoms to recover | Lognormal(14,2.4) | Verity et al. [10] |
| Duration for critical symptoms to death | [mean=5.1 days, std=1.7 days] | Verity et al. [10] |
| **Other model assumptions** |  |  |
| Transmission rate | Calibrated parameter to fit epidemic data |  |
| Relative change in transmission risk when asymptomatic | 0.5 | Assumption |
| Proportion undiagnosed in initial epidemic wave | 40% | Assumption |
| Future testing numbers | 10,000 per day | Assumption based on recent testing blitz in Victoria |
| Sensitivity of test | 70% | Expert opinion |
| Days between having a test and getting result | 1 day | Based on current turnaround time for tests |
| Relative probability of symptomatic people being tested, compared to others | 100 | Assumption based on symptomatic testing policies |

**Table S2:** Age-specific susceptibility, disease progression and mortality risks.

|  | 0-9 | 10-19 | 20-29 | 30-39 | 40-49 | 50-59 | 60-69 | 70-79 | 80+ | Sources |
| --- | --- | --- | --- | --- | --- | --- | --- | --- | --- | --- |
| Relative susceptibility | 0.34 | 0.67 | 1.00 | 1.00 | 1.00 | 1.00 | 1.00 | 1.24 | 1.47 | Zhang et al. [11] |
| Prob[symptomatic] | 0.50 | 0.55 | 0.60 | 0.65 | 0.70 | 0.75 | 0.80 | 0.85 | 0.90 | Assumption |
| Prob[severe] | 0.00004 | 0.00040 | 0.01100 | 0.03400 | 0.04300 | 0.08200 | 0.11800 | 0.16600 | 0.18400 | Verity et al. [10]; CDC [12]. |
| Prob[critical] | 0.0004 | 0.00011 | 0.0005 | 0.00123 | 0.00214 | 0.008 | 0.0275 | 0.06 | 0.10333 | CDC [12] |
| Prob[death] | 0.00002 | 0.00006 | 0.00030 | 0.00080 | 0.00150 | 0.00600 | 0.02200 | 0.05100 | 0.09300 | Verity et al. [10]<br>Ferguson et al. [12, 13] CDC |

### APPENDIX D: Behavioural and contact network parameters for Victoria

The parameters in this appendix were obtained from the literature where available, or through a modified Delphi process where studies were not available (a Delphi process modified to be possible during the COVID-19 pandemic). A group of 12 experts (a mixture of modellers, epidemiologists, qualitative researchers and social network researchers) were invited to participate. A video conference was held where they were introduced to the model and the interpretation of parameters, and participants were asked to make independent estimates of unknown parameters following the conference. Estimates were then collated by the study team, and the median and range of each parameter was extracted. A follow-up video conference was held where the panel discussed the results, uncertainties and were offered an opportunity to update any parameters.

#### Population subsets

Each contact network only applies to a subset of the model population; because not everyone participates in each activity, or attends each location, only a subset are able to be infected at these places or during these activities. The subset of the population that each network applies to is defined as a percentage of a given age range.

**Table S3:**
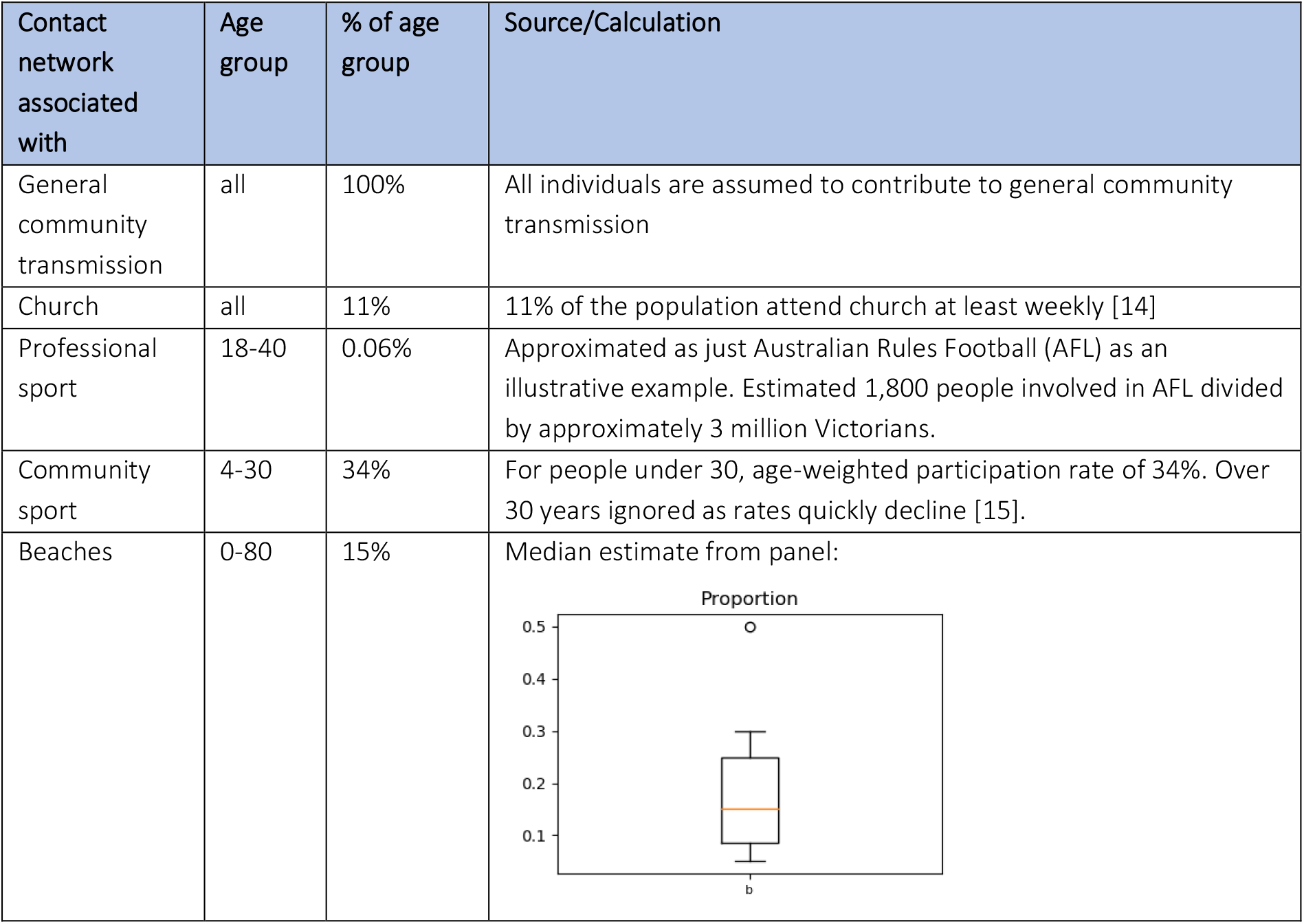
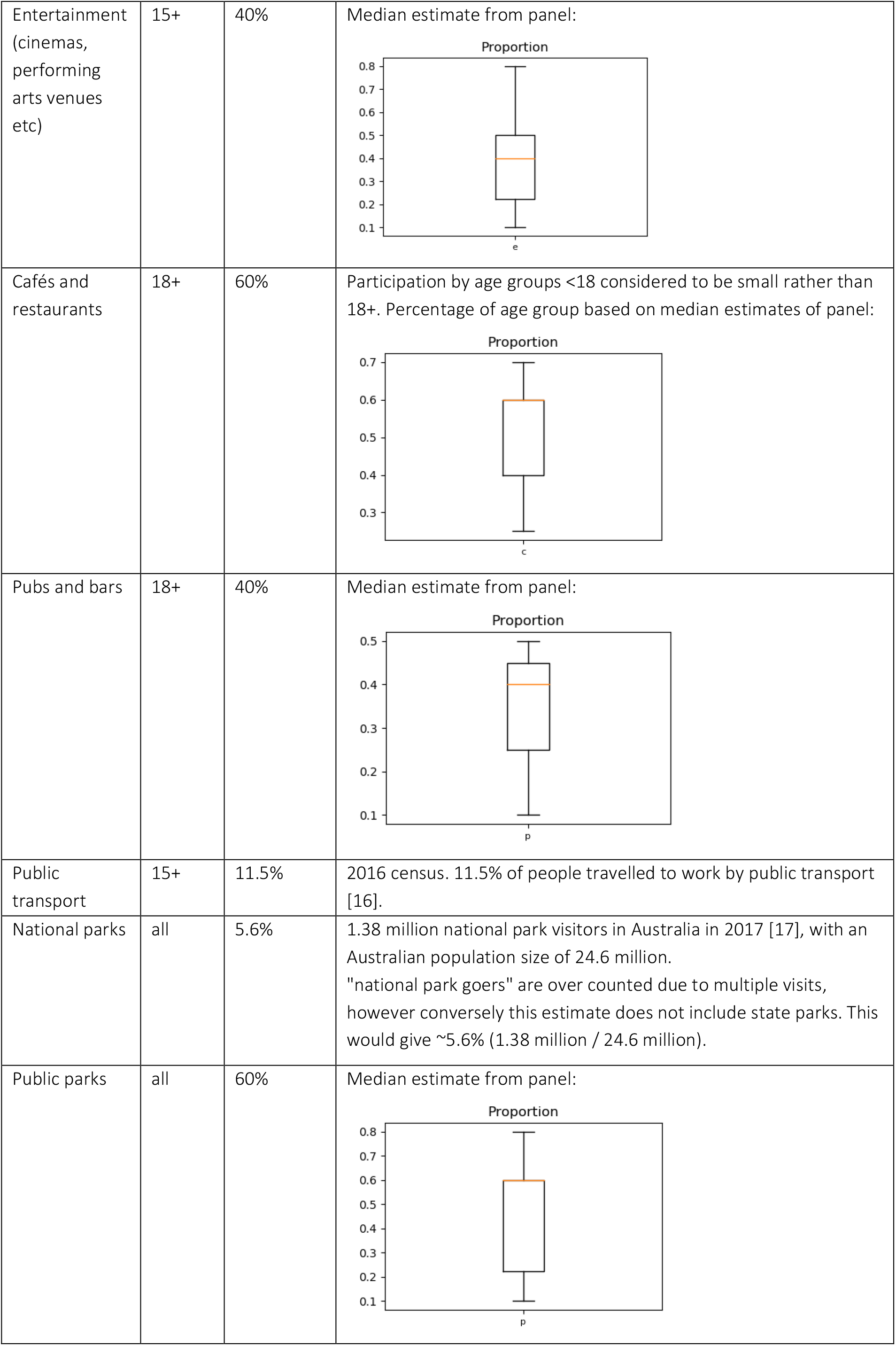
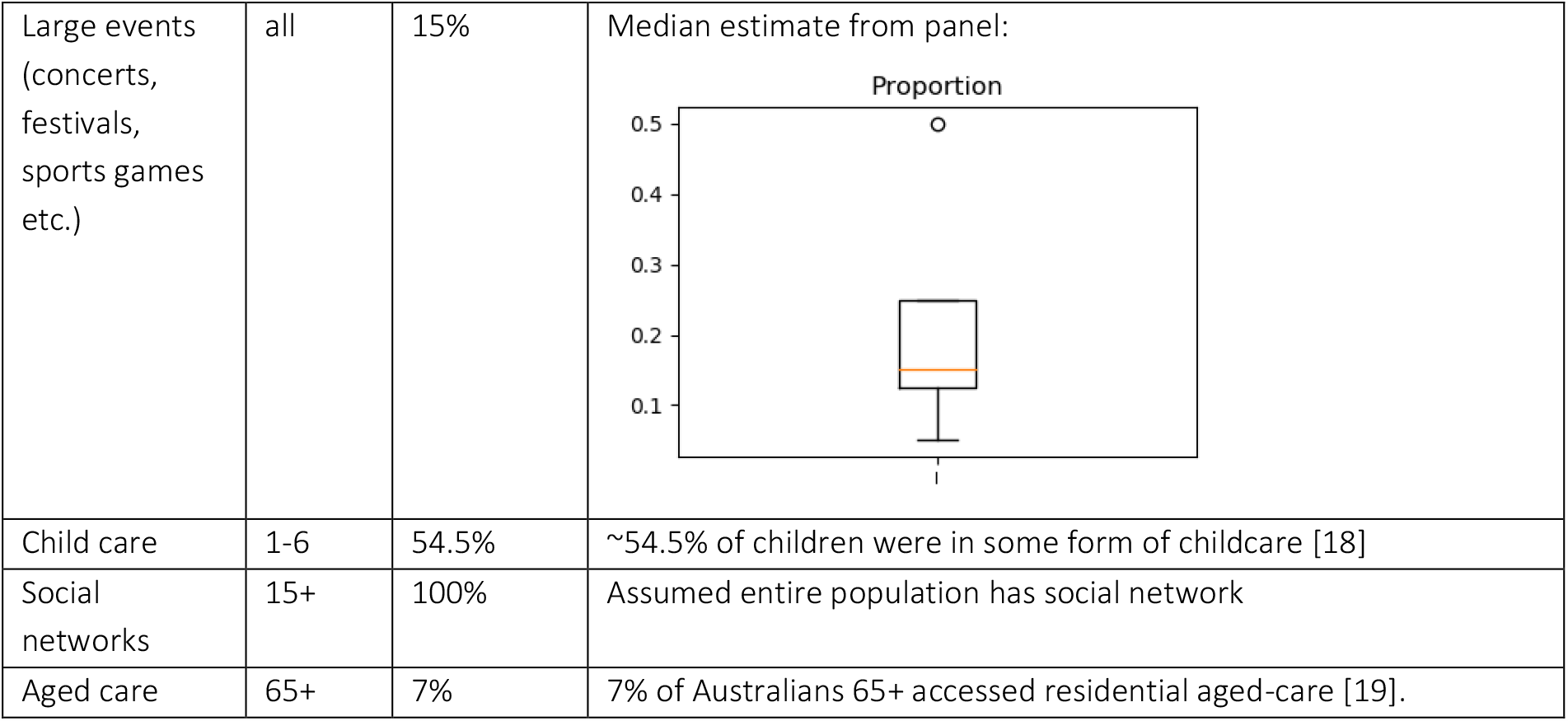
population subsets included in each contact network.

| Contact network associated with | Age group | % of age group | Source/Calculation |
| --- | --- | --- | --- |
| General community transmission | all | 100% | All individuals are assumed to contribute to general community transmission |
| Church | all | 11% | 11% of the population attend church at least weekly [14] |
| Professional sport | 18-40 | 0.06% | Approximated as just Australian Rules Football (AFL) as an illustrative example. Estimated 1,800 people involved in AFL divided by approximately 3 million Victorians. |
| Community sport | 4-30 | 34% | For people under 30, age-weighted participation rate of 34%. Over 30 years ignored as rates quickly decline [15]. |
| Beaches | 0-80 | 15% | Median estimate from panel: |
| Entertainment<br>(cinemas,<br>performing<br>arts venues<br>etc) | 15+ | 40% | Median estimate from panel: |
| Cafés and<br>restaurants | 18+ | 60% | Participation by age groups <18 considered to be small rather than 18+. Percentage of age group based on median estimates of panel: |
| Pubs and bars | 18+ | 40% | Median estimate from panel: |
| Public<br>transport | 15+ | 11.5% | 2016 census. 11.5% of people travelled to work by public transport [16]. |
| National parks | all | 5.6% | 1.38 million national park visitors in Australia in 2017 [17], with an Australian population size of 24.6 million.<br>"national park goers" are over counted due to multiple visits, however conversely this estimate does not include state parks. This would give ~5.6% (1.38 million / 24.6 million). |
| Public parks | all | 60% | Median estimate from panel: |
| Large events<br>(concerts,<br>festivals,<br>sports games<br>etc.) | all | 15% | Median estimate from panel: |
| Child care | 1-6 | 54.5% | ~54.5% of children were in some form of childcare [18] |
| Social<br>networks | 15+ | 100% | Assumed entire population has social network |
| Aged care | 65+ | 7% | 7% of Australians 65+ accessed residential aged-care [19]. |

#### Network structure and size

Each network can have a different structure, with people either being connected to their contacts randomly (“random”) or people being grouped into disconnected clusters (“clustered”, e.g. schools, where the network consists of disjoint classrooms, with students in each classroom connected to one another). The differences between a random and clustered network are illustrated in Figure S4.

Each person in the model has a specified number of contacts in each network layer. The epidemiological definition of a contact between two people is used, where a contact is defined as having a 15-minute face-to-face conversation, or spending one hour or more in a room together. For those who have a non-zero number of contacts in a particular network (i.e. they are inside the applicable age range and randomly-selected population fraction defined in Table S3), if the contact network is “random” type, then their number of contacts is drawn from a Poisson distribution with mean as per Table S4. If the contact network is “clustered”, then the size of each cluster is drawn from a Poisson distribution with mean as per Table S4.

Networks can also be time-varying or not. For example, contact networks for public spaces (e.g. public transport) are regenerated each day, to simulate once-off mixing, compared to work networks in which specific individuals remain connected to one another.

**Table S4:**
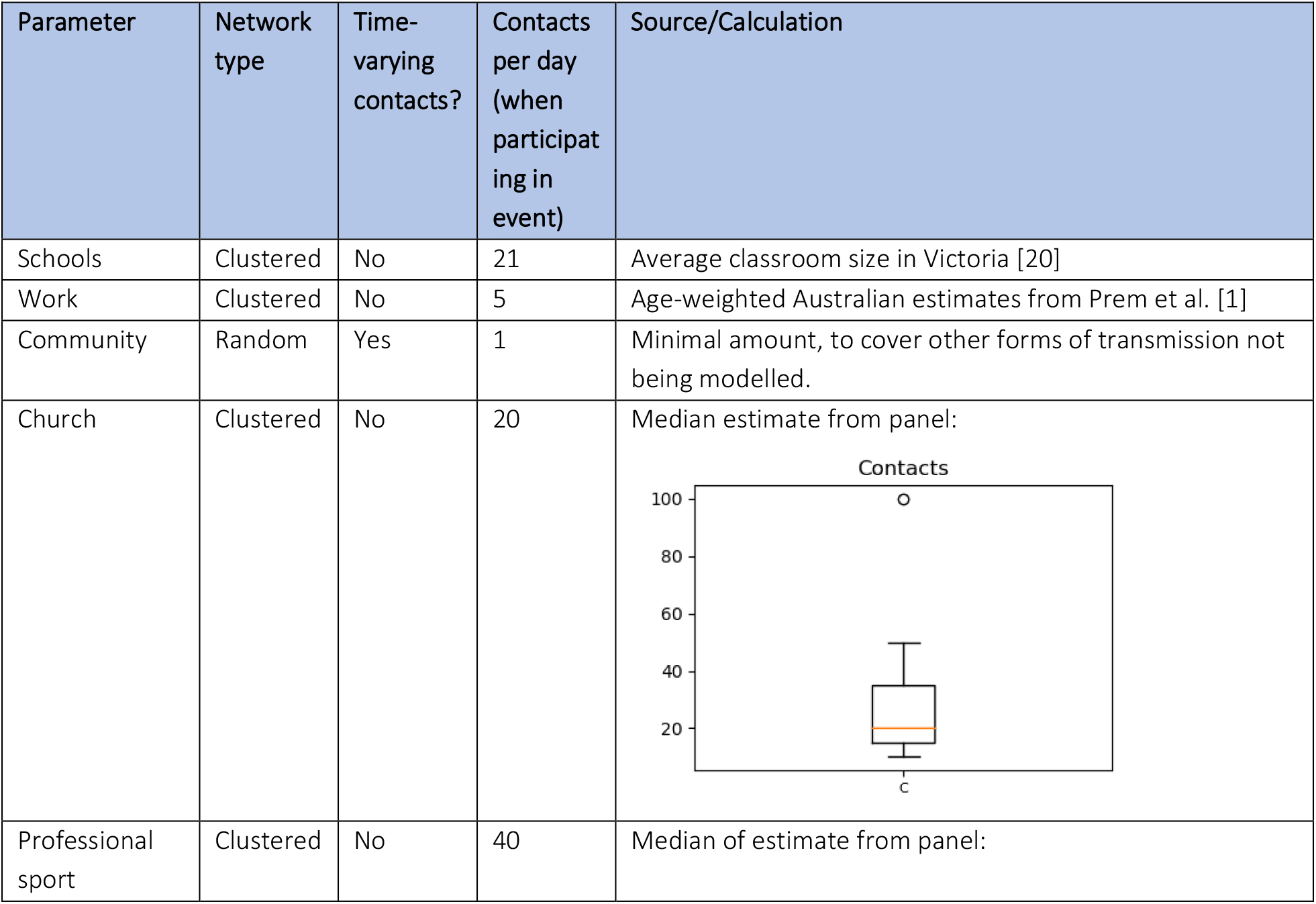
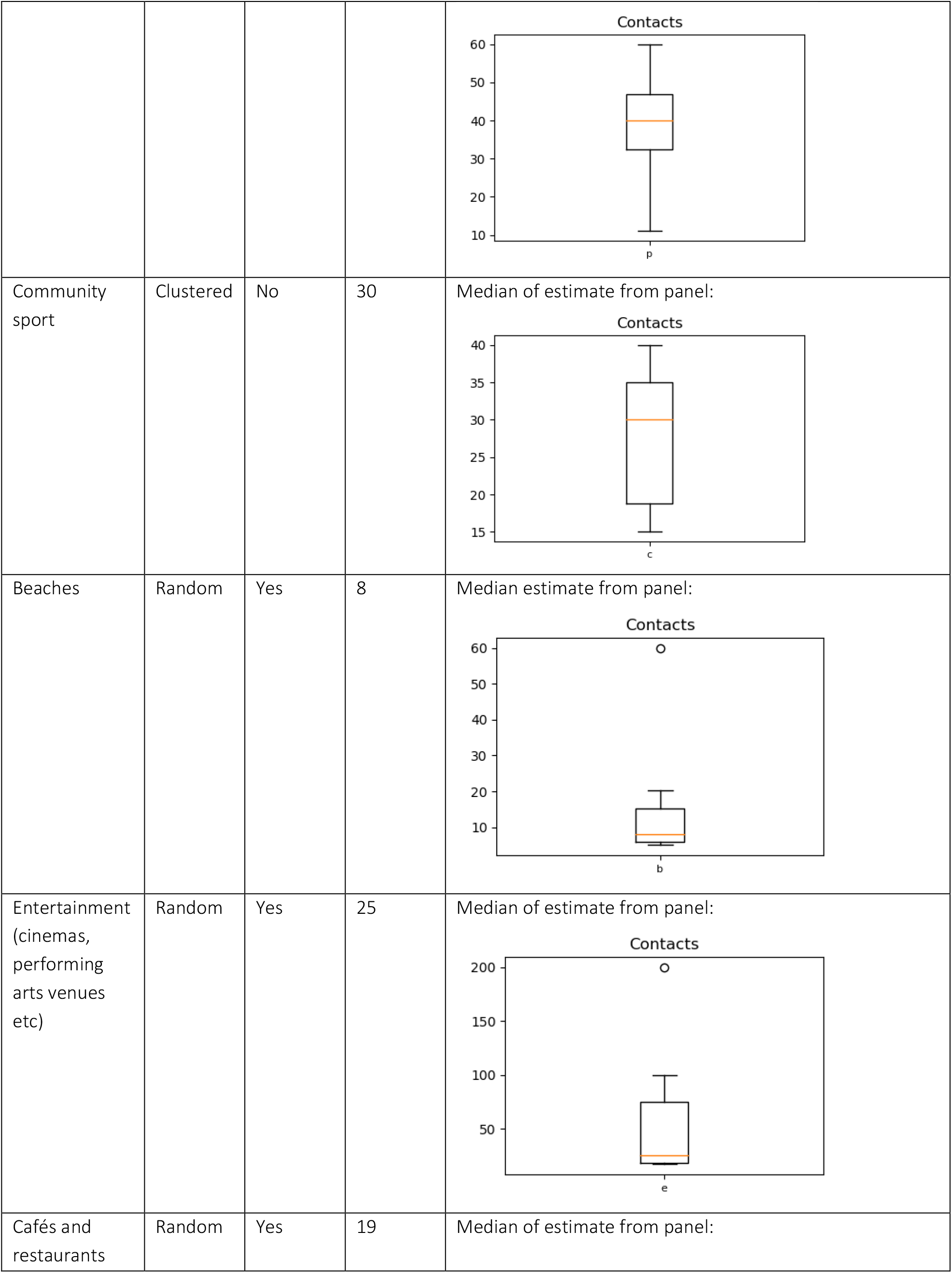
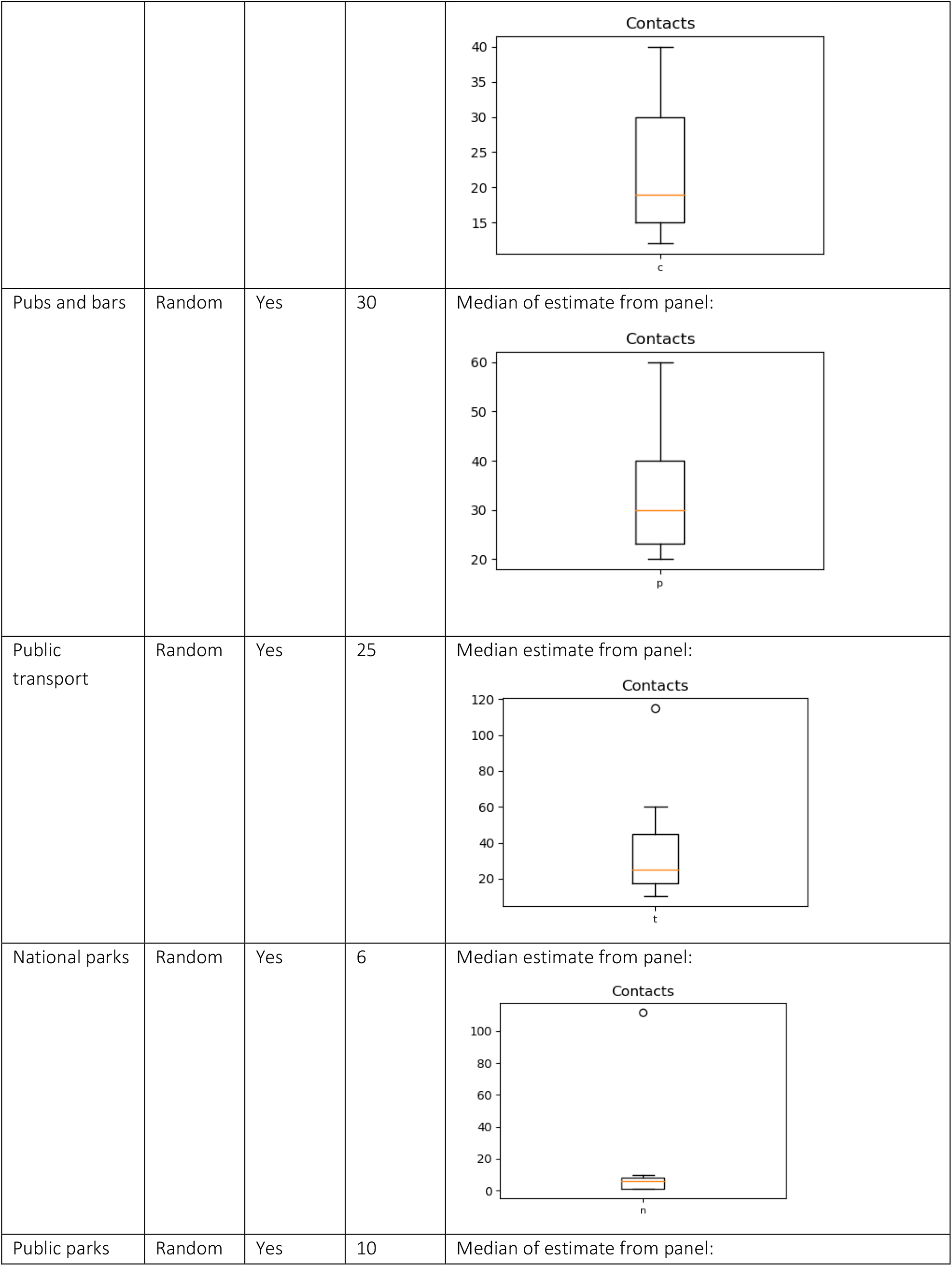
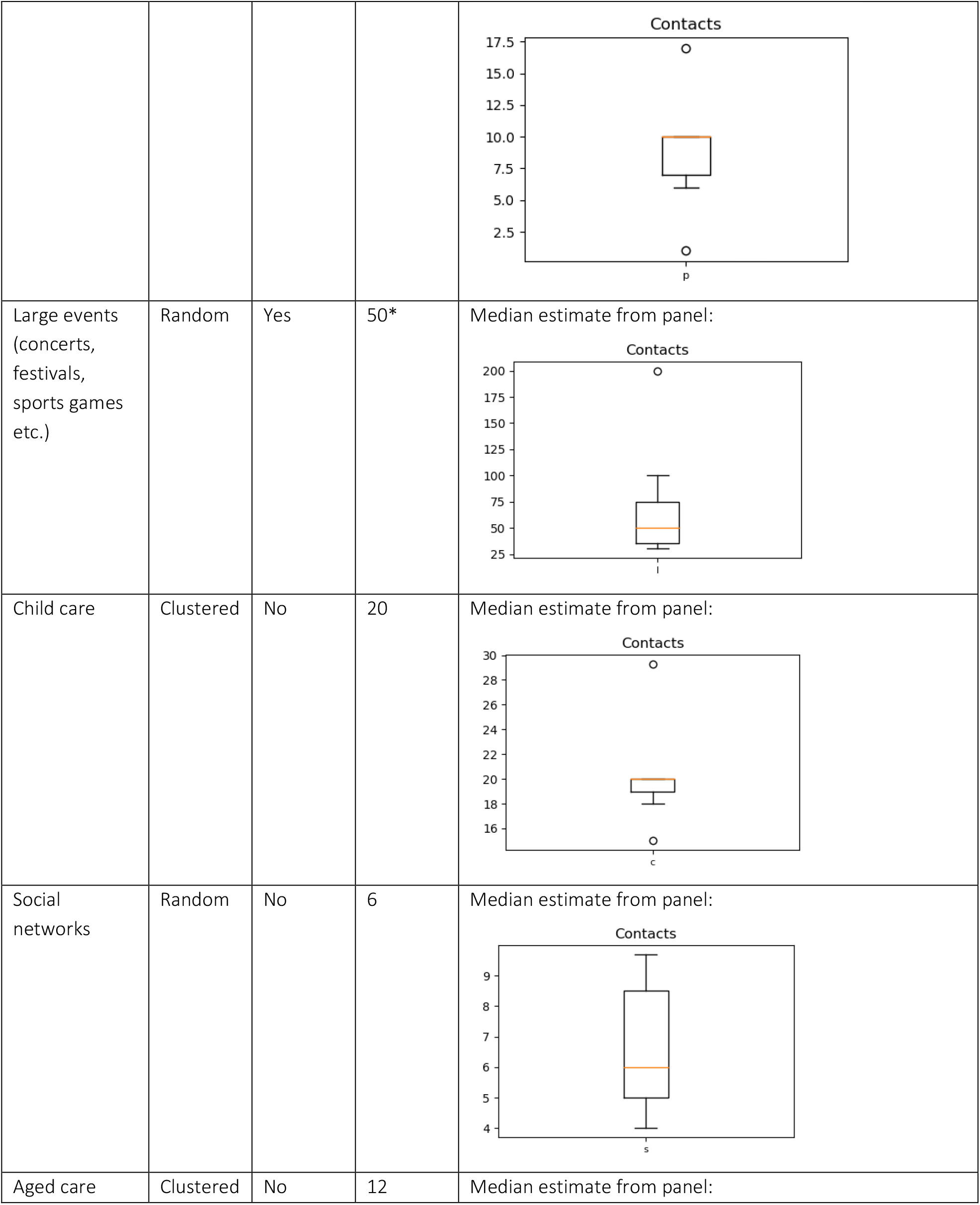
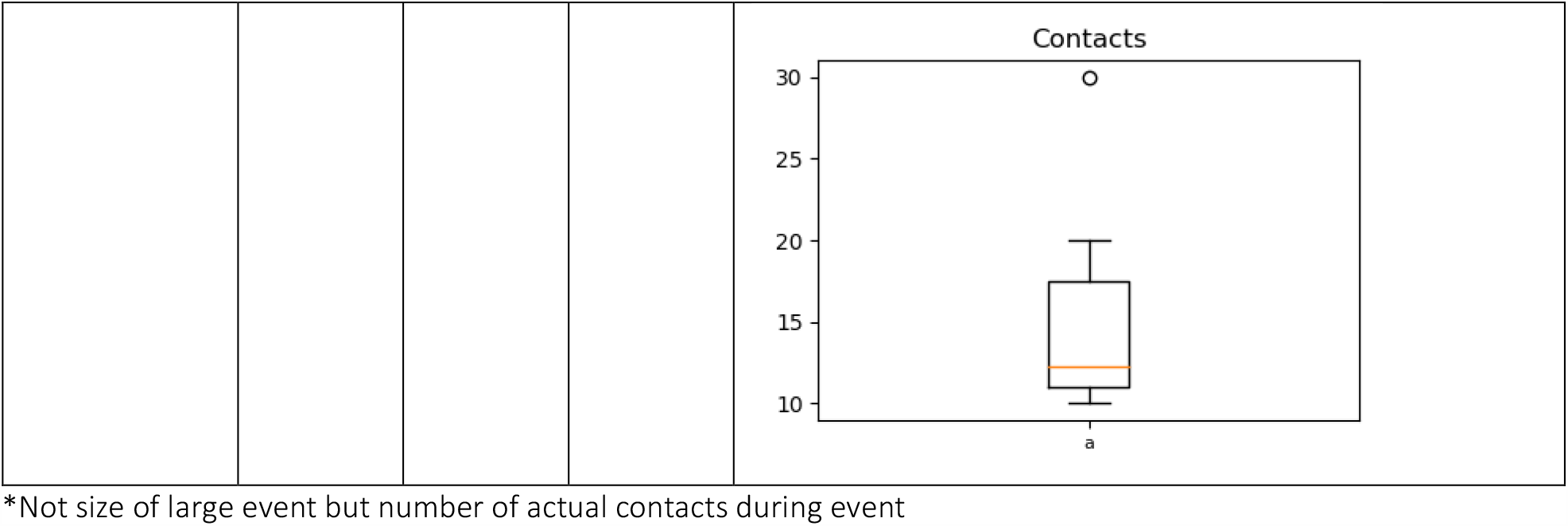
Average number of contacts per person in settings or during activities.

#### Relative transmissibility of contact networks

Transmission of COVID-19 is likely to be highly variable depending on network. As well as an overall daily risk of transmission per contact (the calibration parameter for the model), the risk of transmission per contact per day is different for each network. Table S5 shows these estimated differences relative to the transmission risk per contact per day within households.

**Table S5:**
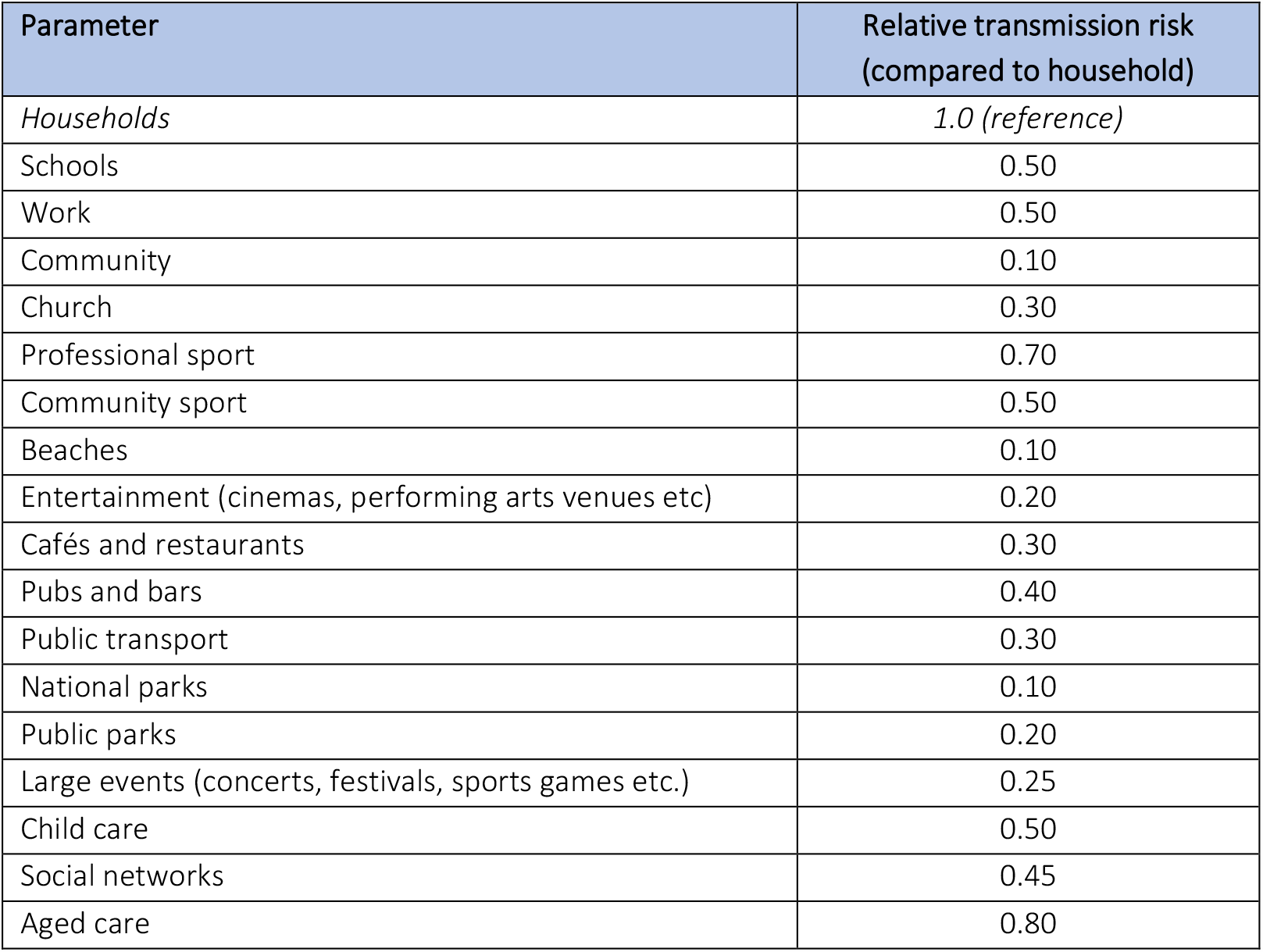
Relative risk of transmission through a contact, compared to a household contact. No studies were available for these parameters, meaning that they were all are based on the median of the expert panel’s estimates shown in Figure S9 below.

| Parameter | Relative transmission risk<br>(compared to household) |
| --- | --- |
| *Households* | *1.0 (reference)* |
| Schools | 0.50 |
| Work | 0.50 |
| Community | 0.10 |
| Church | 0.30 |
| Professional sport | 0.70 |
| Community sport | 0.50 |
| Beaches | 0.10 |
| Entertainment (cinemas, performing arts venues etc) | 0.20 |
| Cafés and restaurants | 0.30 |
| Pubs and bars | 0.40 |
| Public transport | 0.30 |
| National parks | 0.10 |
| Public parks | 0.20 |
| Large events (concerts, festivals, sports games etc.) | 0.25 |
| Child care | 0.50 |
| Social networks | 0.45 |
| Aged care | 0.80 |

**Figure S9:**
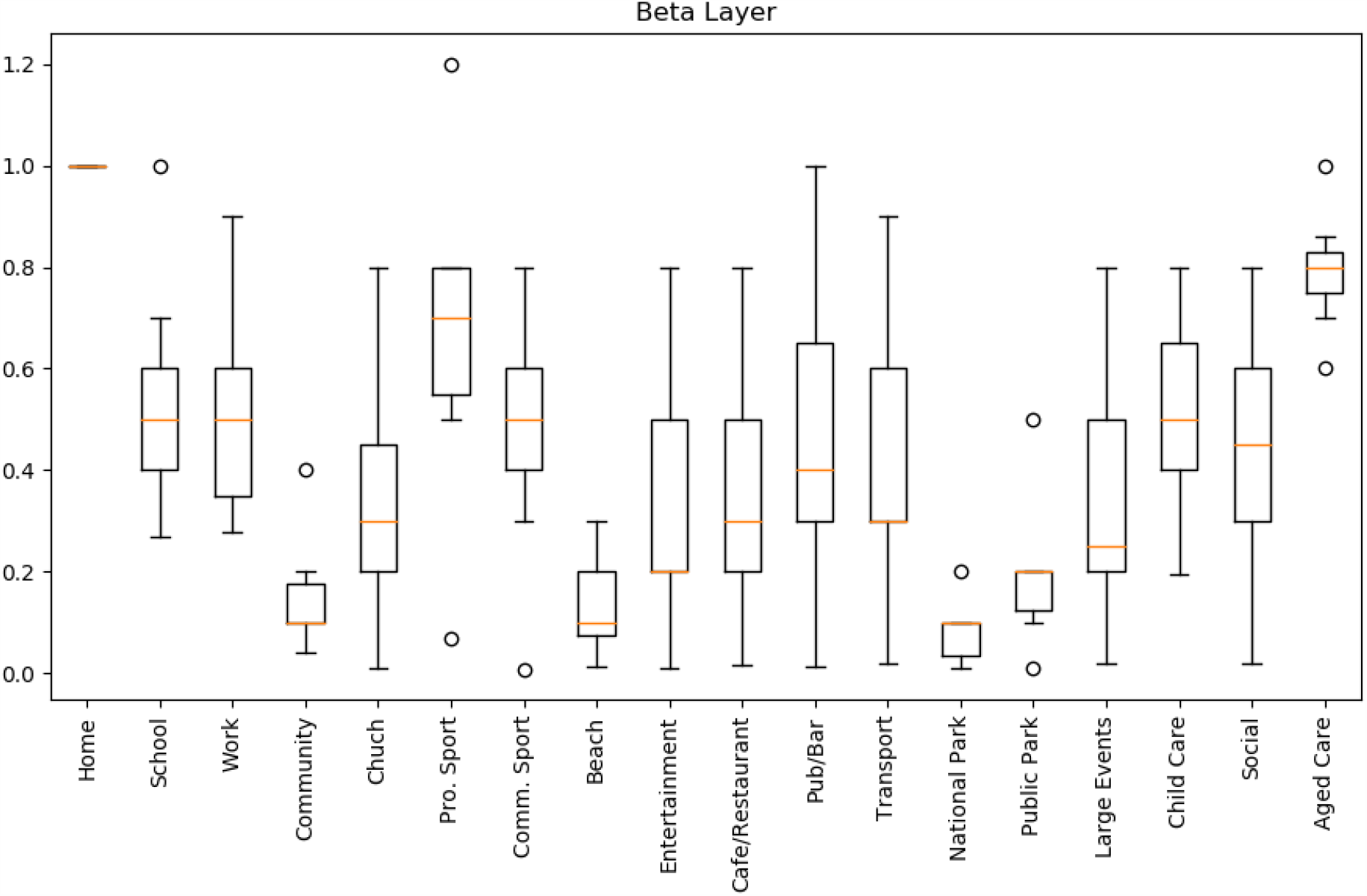
Expert panel estimates for the risk of transmission in each contact network, relative to household contacts.

#### Event frequency

People may not typically interact with the activities and public spaces corresponding to each network on a daily frequency; for example, community sport might be played once per week. The model currently does not include simulation of each activity with different frequencies, and so the impact of this was approximated by reducing the relative transmission risk in each contact network.

The relative transmissibility (Table S5) was divided by the activity frequencies/365 to develop a proxy for per-day transmission risk.

**Table S6:**
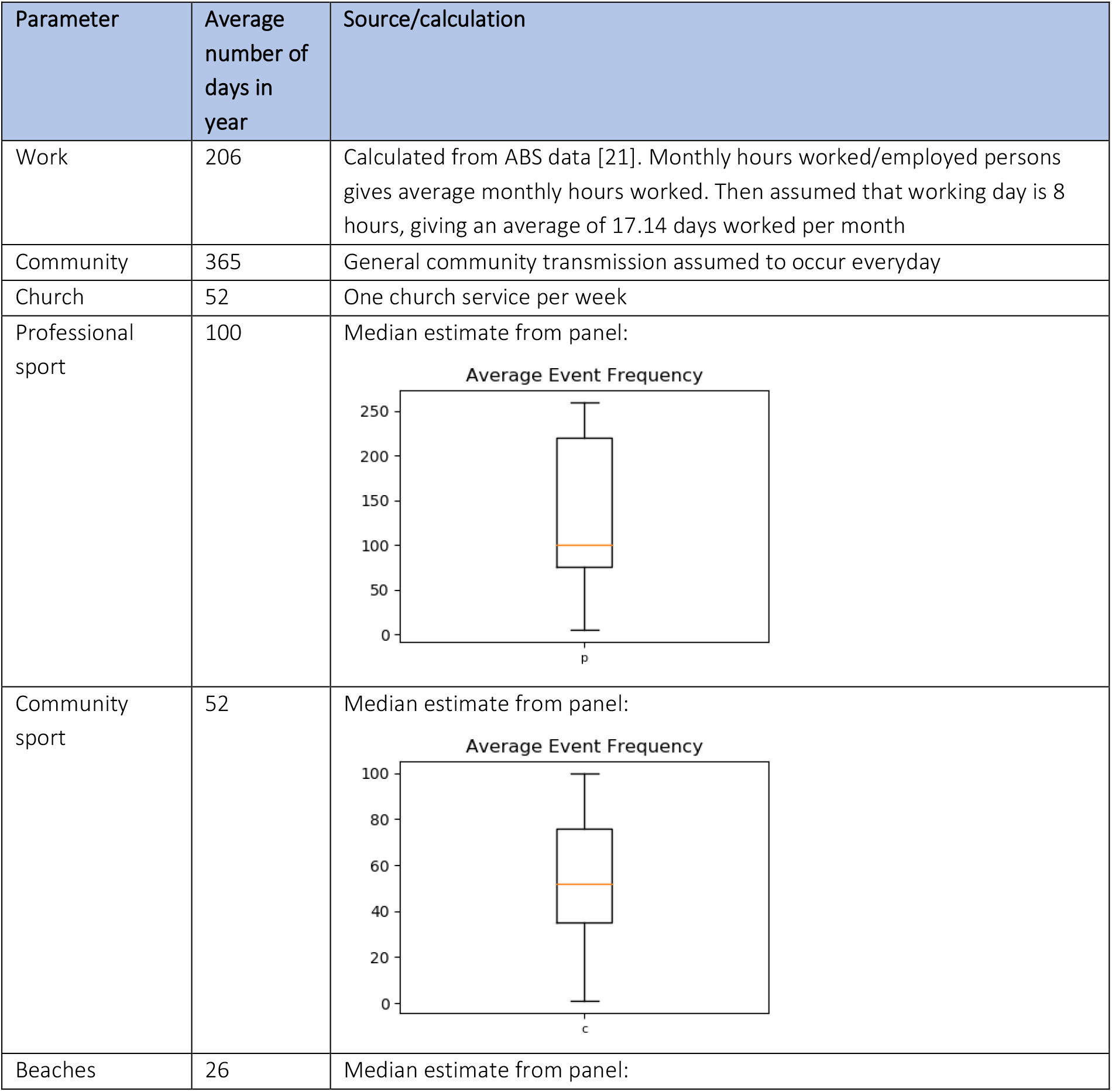
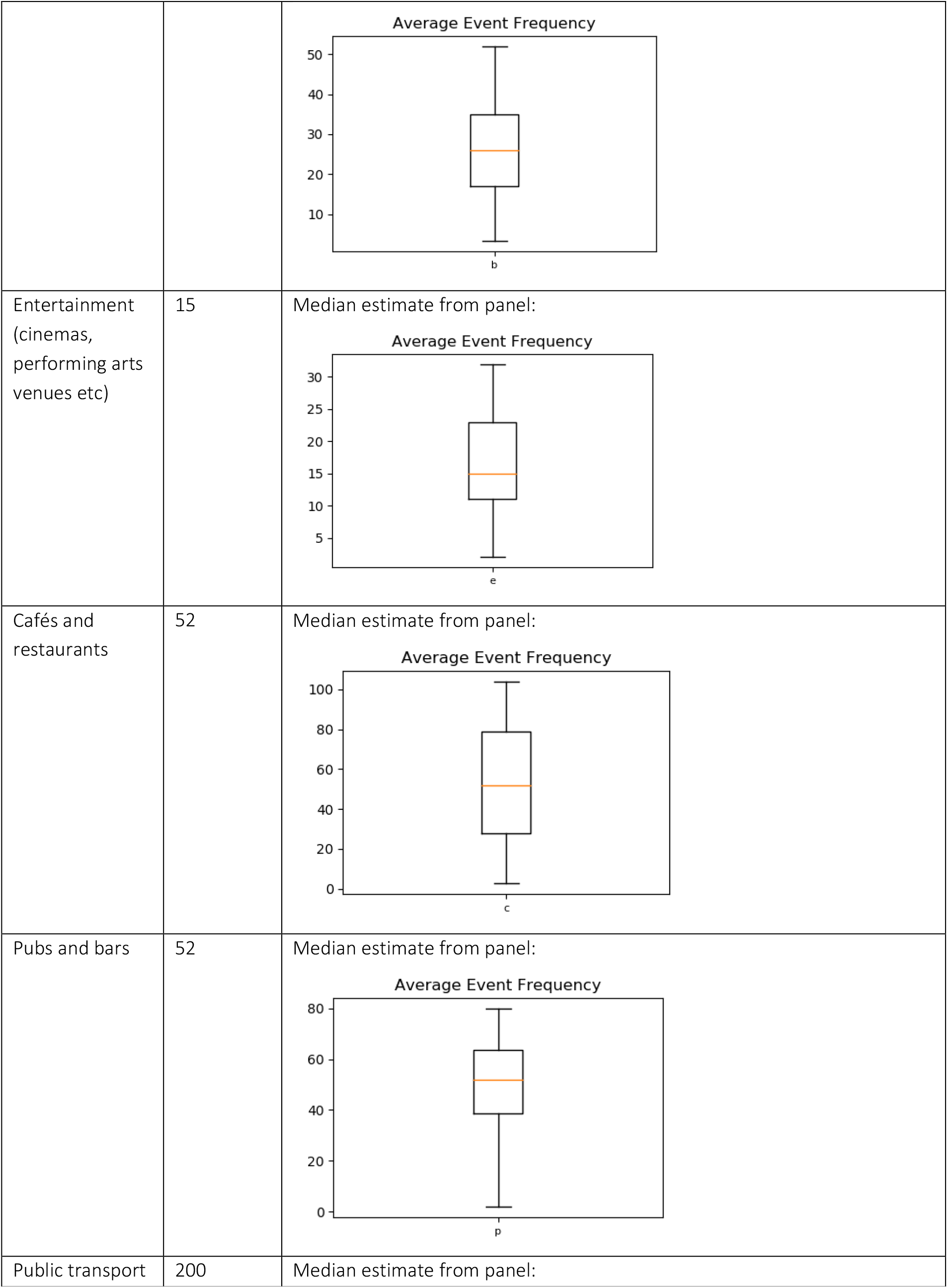
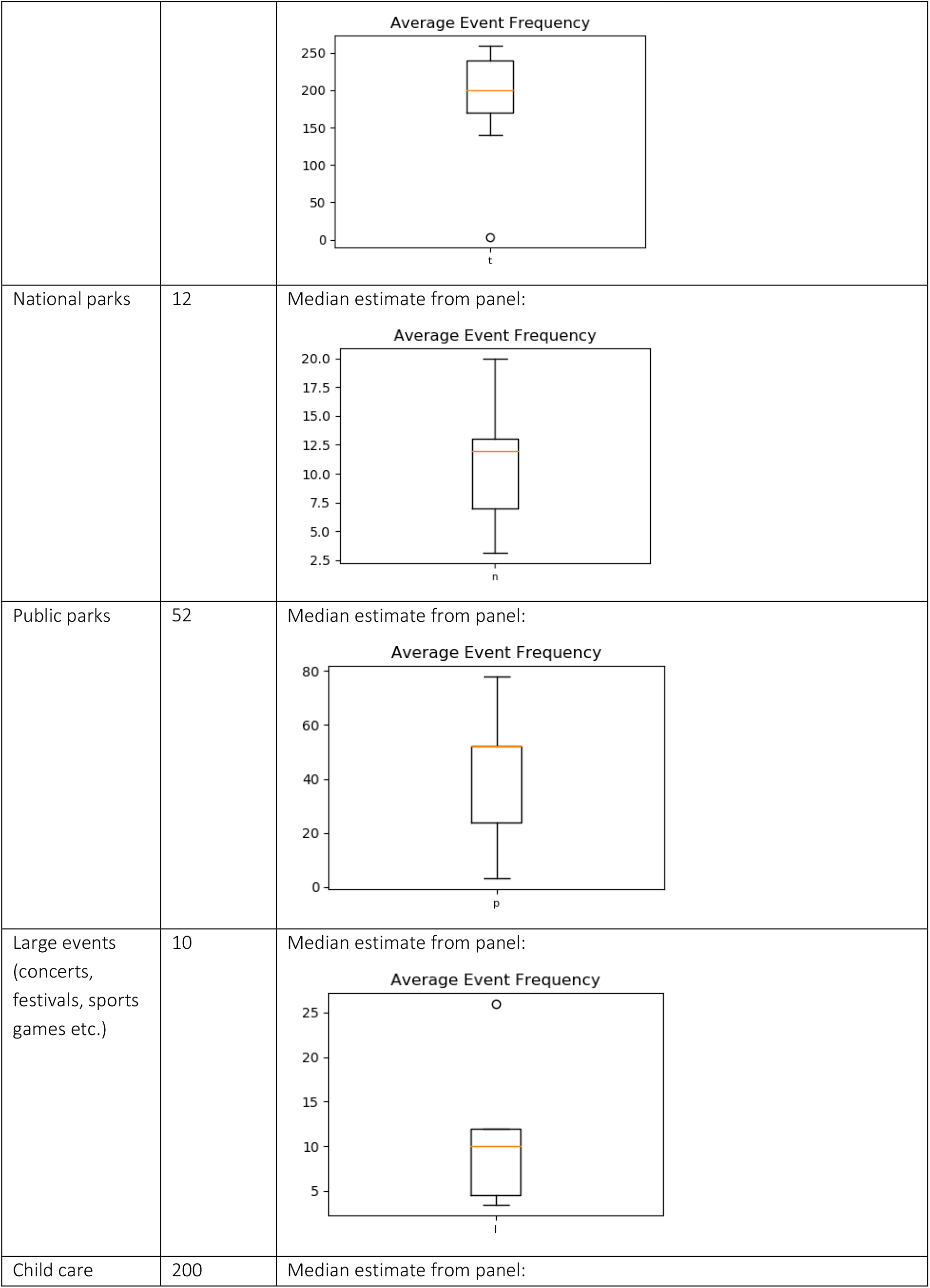
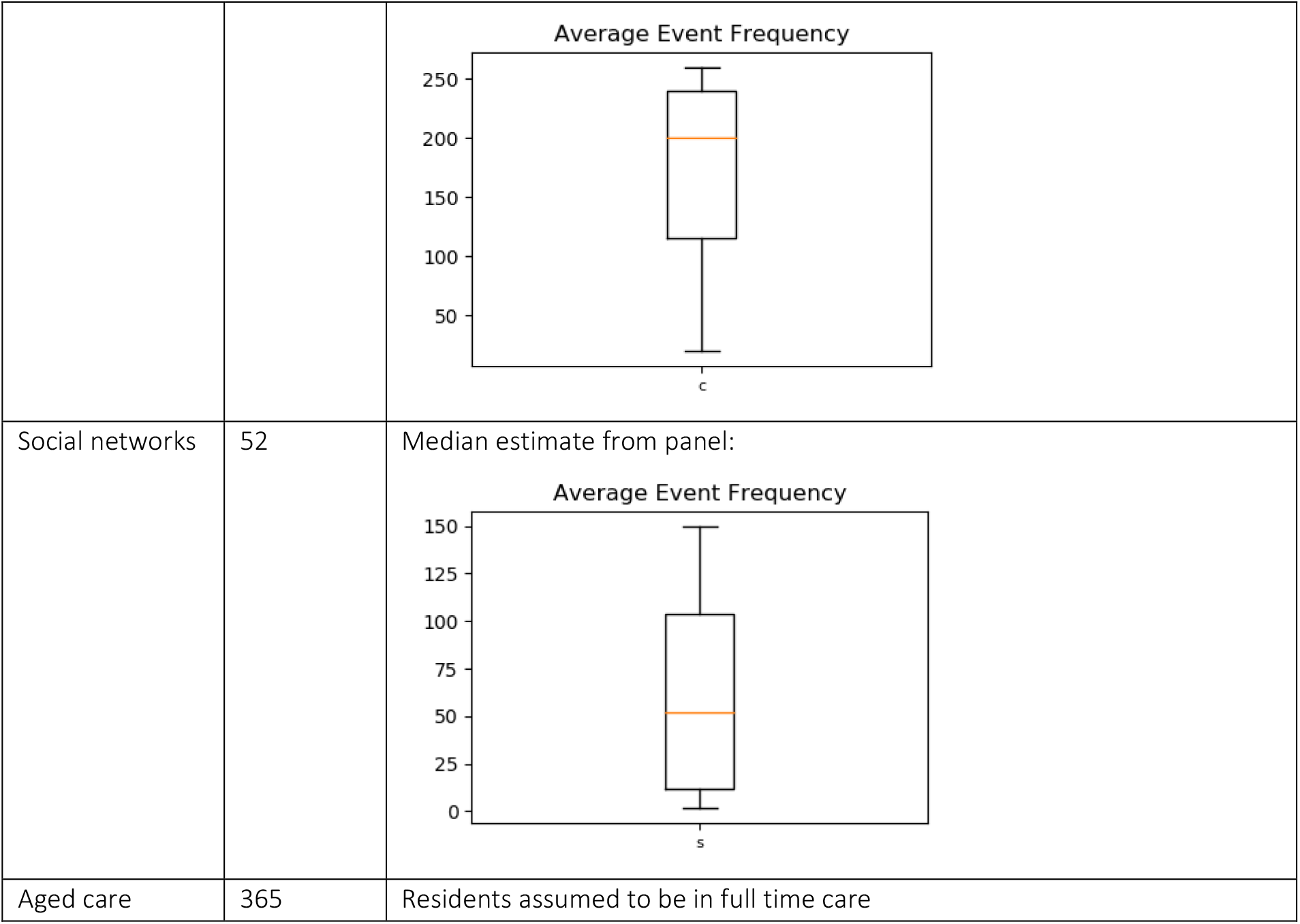
Average Event Frequency.

| Parameter | Average number of days in year | Source/calculation |
| --- | --- | --- |
| Work | 206 | Calculated from ABS data [21]. Monthly hours worked/employed persons gives average monthly hours worked. Then assumed that working day is 8 hours, giving an average of 17.14 days worked per month |
| Community | 365 | General community transmission assumed to occur everyday |
| Church | 52 | One church service per week |
| Professional sport | 100 | Median estimate from panel: |
| Community sport | 52 | Median estimate from panel: |
| Beaches | 26 | Median estimate from panel: |
|  |  | Average Event Frequency<br>b |
| Entertainment<br>(cinemas,<br>performing arts<br>venues etc) | 15 | Median estimate from panel:<br>Average Event Frequency<br>e |
| Cafés and<br>restaurants | 52 | Median estimate from panel:<br>Average Event Frequency<br>c |
| Pubs and bars | 52 | Median estimate from panel:<br>Average Event Frequency<br>p |
| Public transport | 200 | Median estimate from panel: |
|  |  | Average Event Frequency<br>t |
| National parks | 12 | Median estimate from panel:<br>Average Event Frequency<br>n |
| Public parks | 52 | Median estimate from panel:<br>Average Event Frequency<br>p |
| Large events<br>(concerts,<br>festivals, sports<br>games etc.) | 10 | Median estimate from panel:<br>Average Event Frequency<br>l |
| Child care | 200 | Median estimate from panel: |
|  |  | **Average Event Frequency**<br>c |
| Social networks | 52 | Median estimate from panel:<br>**Average Event Frequency**<br>s |
| Aged care | 365 | Residents assumed to be in full time care |

#### Quarantine and contact tracing

People who are asked to self-isolate are likely to change their behaviour in ways that reduce their likelihood of transmission through different contact networks. For people in quarantine, their relative transmissibility in each contact network (Table S5) is reduced by the factors shown in Table S7. For example, quarantine is modelled to have no impact on household transmission, to completely stop workplace and school transmission, and reduce (but not stop) other forms of community transmission due to imperfect adherence.

When a person is diagnosed, there is a probability of tracing the people they are connected to in different contact networks, and an associated time to trace them. For example, we assume that household members would be notified on the day of diagnosis, while workplace contacts would have a 70% chance of being traced within 2 days.

The effectiveness of quarantine, contact tracing probabilities and tracing time were estimated from the expert panel.

**Table S7:** Effectiveness of quarantine and contact tracing on different contact networks. No studies were available for these parameters, meaning that they were all are based on the median of the expert panel’s estimates.

| Parameter | Quarantine effectiveness | Probability of successful contact tracing | Average time to trace contact |
| --- | --- | --- | --- |
| Households | 1.00 | 1.00 | 1 |
| Schools | 0.01 | 0.95 | 2 |
| Work | 0.10 | 0.80 | 2 |
| Community | 0.20 | 0 | N/A |
| Church | 0.01 | 0.50 | 5 |
| Professional sport | 0.00 | 0.80 | 3 |
| Community sport | 0.00 | 0.50 | 3 |
| Beaches | 0.00 | 0 | N/A |
| Entertainment (cinemas, performing arts venues etc) | 0.00 | 0 | N/A |
| Cafés and restaurants | 0.00 | 0 | N/A |
| Pubs and bars | 0.00 | 0 | N/A |
| Public transport | 0.01 | 0 | N/A |
| National parks | 0.00 | 0 | N/A |
| Public parks | 0.00 | 0 | N/A |
| Large events (concerts, festivals, sports games etc.) | 0.00 | 0 | N/A |
| Child care | 0.01 | 0.95 | 2 |
| Social networks | 0.00 | 0.90 | 3 |
| Aged care | 0.20 | 0.95 | 2 |

#### Intervention effectiveness

There were no studies available to estimate the impact of policy changes on each network. However, for many polices, the impact is based on turning on / off transmission within a particular network, and so the impact is derived from the network properties in Tables S3–S6.

For some policies, there are logical impacts that extend beyond their specific network; for example, if non-essential work is cancelled, then the transmission risk on public transport would be expected to decrease. For these auxiliary effects, the actual impact size is unknown, and so has been estimated by the panel of experts.

**Table S8:** Impact of policies. Data were not available to inform changes in transmission due to different policies. All estimates are based on median values reported

| Description | Parameter changes (compared to pre-COVID time) |
| --- | --- |
| Physical distancing communication and enforcement | 86% decrease in overall beta* |
| Physical distancing communication and enforcement relaxed a bit (when restrictions begin to be lifted) | When physical distancing is relaxed, overall hygiene and physical distancing benefits are reduced by 75% (from 86% reduction (see above) to only a 35% reduction) |
| Beaches closed | 0 transmission risk in beach network |
| Beaches restricted to groups of 2 | 80% decrease in transmission risk within beach network |
| Beaches restricted to groups of <10 | 40% decrease in transmission risk within beach network |
| National and state parks closed | 0 transmission risk in national park network |
| Churches / places of worship closed | 0 transmission risk in church network |
| Churches / places of worship implementing 4 sq m rule | 40% decrease in transmission risk within church network |
| Cafes and restaurants take-away only | • 10% increase in transmission risk at home<br>• 0 transmission risk in café_restaurant network |
| Cafes and restaurants implementing 4 sq m physical distancing rule | 50% decrease in transmission risk within café_restaurant network |
| Pubs and bars closed | • 10% increase in transmission risk at home<br>• 0 transmission risk in pub_bar network |
| Pubs and bars implementing 4 sq m physical distancing rule | 40% decrease in transmission risk within pub_bar network |
| Outdoor settings restricted to <2 people | • 20% increase in transmission risk at home<br>• 30% decrease in general community transmission risk<br>• 30% decrease in transmission risk in transport network<br>• 0 transmission risk in entertainment network<br>• 0 transmission risk in national park network |
|  | • 60% decrease in transmission risk in public park network<br>• 0 transmission risk in large event network<br>• 70% decrease in transmission risk in social networks |
| Outdoor settings restricted to <10 people | • 5% increase in transmission risk at home<br>• 20% decrease in transmission risk in general community network<br>• 0 transmission risk in entertainment network<br>• 30% decrease in transmission risk in transport network<br>• 30% decrease in in transmission risk in public park network<br>• 0 transmission risk in large event network |
| Outdoor settings restricted to <200 people | • 20% decrease in transmission risk in transport network<br>• 0 transmission risk in large event network |
| Professional sports cancelled for players (crowds are different policy) | 0 transmission risk in pSport network |
| Community sports cancelled | 0 transmission risk in cSport network |
| Child care closed | 0 transmission risk in child_care network |
| Schools closed | • 50% decrease in transmission risk in school network<br>• 90% of children removed from school network |
| Non-essential retail outlets, including shopping centres closed | • 30% decrease in transmission risk in general community network<br>• 5% of workers are removed from work network |
| Cinemas, performing arts venues etc. closed | 0 transmission risk in entertainment network |
| concerts, festivals, sports games etc. | 0 transmission risk in large event network |
| Non-essential work closed | • 33% reduction in transmission risk on public transport<br>• 20% of workers are removed from work network |
| Non-COVID-19 health services closed | 5% of workers are removed from work network |
| Travel across state borders allowed and increased domestic travel | imported infections increases to 5 per day |
| social catch ups with <10 people banned | 0 transmission risk in social network |
| Enhanced screening and distancing within age care facilities | 0 transmission risk in aged care network |

\* From flutracker, 0.2% fever and cough prevalence compared to ~1.4% the same time last year --> 86% reduction [22].

### APPENDIX E: Policy changed to be simulated in the model

Interventions can be modelled by changing parameters dynamically throughout a simulation. At any time point in a simulation, parameters can be varied to:

- Change the number of imported infections (from other Australian jurisdictions or internationally)
- Change the number of tests per day
- Change adherence to quarantine after diagnosis
- Scale the overall probability of transmission per contact (e.g. due to general hand hygiene)
- Scale the relative transmission risk for specific contact layers (e.g. a policy closing cafes and restaurants would set the transmission risk for the cafe/restaurant network to be zero)
- Remove a proportion of people from a network (e.g. a policy stopping non-essential work would remove some people from the work contact network)
- Change the effectiveness of contact tracing for a particular contact network (e.g. the COVIDSafe app makes contact tracing possible for community transmission only if both the infected and susceptible person have the app)

Policy changes are linked to one or more networks, and can potentially influence the whole population. For example, if non-essential work begins, this would increase the size of the work network, as well as increasing transmissibility in public transport.

Policy scenarios modelled were informed by the COVID-19 public health response and the COVIDSAFE Australia framework [23]. The following are examples of policies that can be simulated:

1. Contact tracing (including the use of COVIDSafe app for different coverages)
2. Communication and enforcement of physical distancing (e.g. signs, advertisements, policing)
3. Cafes and restaurants take-away only
4. Cafes and restaurants implementing 4 square metre rule physical distancing rule
5. Pubs and bars closed
6. Pubs and bars implementing 4 square metre rule physical distancing rule
7. Churches / places of worship closed
8. Churches / places of worship implementing 4 square metre rule physical distancing rule
9. Outdoor settings restricted to <2 people
10. Outdoor settings restricted to <10 people
11. Outdoor settings restricted to <200 people
12. Indoor social catch ups with <10 people banned
13. Community sports
14. Professional sports (for players)
15. Child care closed
16. Schools closed
17. Entertainment venues closed (e.g. cinemas, performing arts)
18. Large events cancelled (e.g. concerts, festivals, sports games)
19. Beaches closed
20. Beaches restricted to groups of 2
21. Beaches restricted to groups of <10
22. National and state parks closed
23. Non-essential retail outlets closed
24. Non-essential work closed
25. Non-COVID-19 health services closed
26. Travel restrictions across state borders

Any set of interventions can be run in combination, or staged according to policy change dates.

## Notes

### Competing Interest Statement

The authors have declared no competing interest.

### Funding Statement

No external funding was received for this study.

### Author Declarations

No ethics were required as all data used for this modelling study were publicly available.

